# Anticipatory nursing work and the co-construction of discharge readiness in coronary care units: A cross-national qualitative study

**DOI:** 10.64898/2026.08.03.26358968

**Authors:** Amineh Rashidi, Melissa Dunham, Emma Kinley, Teofila Bueser, Courtney Glass, Ian Jones, Mark Makokha, Lisa Whitehead, Lisa Newson

**Affiliations:** School of Nursing Curtin School of Nursing Faculty of Health Sciences, Curtin University; University of Melbourne, School of Population and Global Health; Liverpool John Moores University, School of Psychology; King’s College London; Department of Education Western of Australia; School of Nursing and Allied Health, Liverpool John Moores University; South Metropolitan Health Service. Fiona Stanley Hospital, Murdoch, Western Australia; School of Nursing and Midwifery, Edith Cowan University

**Keywords:** Patient discharge, Coronary care units, Nursing practice, Interprofessional collaboration, Transitional care, Anticipatory care, Qualitative research

## Abstract

**Aim:** To conceptualise discharge readiness in CCUs as anticipatory nursing and interprofessional work and to examine how professional roles, organisational systems and service contexts shape discharge across two national settings.

**Design:** Qualitative study utilising reflexive thematic analysis.

**Methods:** Semi-structured interviews and focus groups were conducted with 37 clinicians working in CCUs in Australia and the United Kingdom. The Australian sample comprised nurses, allied health professionals and medical staff; the UK sample comprised nurses only. Data were analysed using reflexive thematic analysis within each dataset, followed by cross-site interpretive integration.

**Results:** Four interrelated themes were generated: (1) anticipatory nursing work initiated at admission and sustained through ongoing assessment and coordination; (2) discharge readiness as a collective accomplishment co-constructed through interprofessional negotiation; (3) documentation as an organisational mechanism that shaped how discharge work was prioritised and enacted; and (4) discharge work enacted within, not separately from, the system and organisational conditions that constrained it. Across both settings, nurses held the coordinative work together, although its expression varied with local organisational scaffolding.

**Conclusion:** Discharge readiness in coronary care is better understood as continuous, anticipatory work than as an endpoint. The findings make visible the distributed and often invisible labour through which discharge readiness is produced and the organisational conditions that support or constrain it. The paper contributes to nursing scholarship on the invisible work of coordination in nursing and offers a conceptual model and practice-informed checklist to support anticipatory discharge work.

**Implications for the profession and/or patient care:** *Impact:* What problem did the study address?
Hospital discharge from coronary care units (CCUs) is commonly framed as a discrete clinical or administrative endpoint.
This event-based conceptualisation obscures the ongoing nursing and interprofessional work undertaken across the admission to sustain discharge readiness.
It is therefore important to examine how professional roles, organisational systems and service context shape discharge across two national settings. What were the main findings?
Reconceptualises discharge readiness in coronary care as continuous, anticipatory nursing work, co-constructed across the admission rather than confirmed at its end.
Draws on a health psychology lens to theorise the cognitive and emotional demands of anticipatory discharge work on individual clinicians.
Presents an empirically grounded conceptual model and a practice-informed clinician checklist derived directly from the findings. Where and on whom will the research have an impact?
This study uses nurses’ voices to offer insights for a holistic understanding of their professional roles, organisational systems in the discharge context. This study offers knowledge relevant to multidisciplinary input and support coordinated and safe discharge planning. Understanding discharge context as continuous anticipatory work than as an endpoint is essential to reinforces the role of the established allied health and community nursing infrastructure. Nurses are pivotal in this process, making enhanced understanding crucial for improving high-quality discharge planning, and opportunities to improve transitional care. No Patient or Public Involvement
This study did not include patient or public involvement in its design, conduct, or reporting. Reporting Method
COREQ and Reflexive Thematic Analysis Reporting Guidelines.

## 1. Introduction

Hospital discharge is a critical transition that influences patient safety, continuity of care and how well people manage their condition once home. In coronary care units (CCUs), where patients are admitted with acute cardiac events, the stakes are particularly high. The timing and quality of discharge influence readmission risk, uptake of secondary prevention and the efficient use of scarce monitored beds (Feltner et al., 2014; Naylor et al., 2011). Despite its importance, discharge is still routinely treated as a discrete event, a documentation task, a medical sign-off, or the moment a patient simply leaves the bed, obscuring the substantial anticipatory clinical work required to produce a safe discharge long before the patient is medically cleared to leave.

Discharge planning and discharge readiness are often used interchangeably in clinical and research literature. Discharge planning typically denotes the formal, structured activities undertaken to prepare a patient for leaving hospital, needs assessment, service arrangement, patient education and documentation, and has been the primary focus of transitional care research and intervention development (Shepperd et al., 2013; Tyler et al., 2023). Discharge readiness, by contrast, refers to the condition of being genuinely prepared for safe discharge: clinically stable, functionally capable, psychosocially supported and practically resourced for the transition home. This paper’s central argument is that discharge readiness is not a fixed state confirmed at a single assessment point but is continuously produced through anticipatory work across the admission, work that is not captured by the concept of discharge planning and is largely invisible within event-based accounts of discharge.

## 2. Background

Cardiovascular disease remains the leading cause of death worldwide (Leach-Kemon, Loeffler, and Islam, 2025), placing sustained pressure on acute services in both Australia and the United Kingdom (UK). Workforce shortages, high bed occupancy and limited community capacity mean that discharge timing is not only a clinical concern but also a major organisational challenge. In response, several transitional care models have been developed, including structured discharge planning, enhanced patient education and post-discharge follow-up programmes such as the Care Transitions Intervention and the Transitional Care Model (Coleman et al., 2006; Naylor et al., 2011). Cardiac-specific systematic reviews indicate that these interventions produce variable effects depending on context (Feltner et al., 2014; Tyler et al., 2023). However, most studies have focused on post-discharge support or the procedural components of discharge planning (Leithaus et al., 2022), with comparatively little qualitative research examining how discharge readiness is produced through the everyday clinical work.

Much of the work that produces a safe discharge in coronary care is performed by nurses and is not visible in formal discharge processes, because discharge rarely occurs as a single decision. Instead, it develops through repeated assessments of clinical stability, interprofessional coordination, negotiation with patients and families, and ongoing risk judgement carried out alongside everyday patient care. This work is shaped by clinical status, patient capability, social support, organisational workflows and time pressures (Naylor et al., 2011). Existing research has highlighted inconsistencies in discharge summaries, fragmented communication between professional groups and gaps in information transfer between hospital and primary care (Hesselink et al., 2012; Kripalani et al., 2007). These findings align with longstanding recognition of the often-invisible organisational labour nurses perform to maintain care coordination and patient safety (Allen, 2014) and suggest that discharge is shaped by team dynamics and broader service contexts well beyond its logistical dimensions.

Australia and the UK share publicly funded healthcare systems and broadly comparable standards of cardiac care, but they differ in organisational structures that shape how discharge is accomplished. The UK National Health Service (NHS) is a centrally directed system supported by national guidance on care transitions (NICE, 2015) and relatively well-developed community nursing infrastructure; however, it is frequently constrained by social care capacity, which can delay discharge even when patients are clinically stable (Glasby et al., 2021). In contrast, the Australian public health system is federated across states and territories, characterised by greater workforce diversification, including embedded clinical pharmacists with direct discharge responsibilities, alongside significant geographic challenges affecting medication access in regional and rural areas (ACSQHC, 2017). These contextual differences mean that the same clinical task is enacted through partly overlapping yet distinct organisational routines, and comparing the two setting brings shared anticipatory processes into focus while drawing out the influence of local organisational scaffolding.

The comparison is best understood as interpretive contrast rather than equivalence testing; differences in professional composition and data collection format across the two datasets are addressed in the Methods. The intent is not to rank the two systems but to use the contrast to illustrate how organisational context shapes what discharge work looks like in practice. This study therefore aims to conceptualise discharge readiness in coronary care units as anticipatory nursing and interprofessional work, and to examine how professional roles, organisational systems and service context shape discharge across two national settings. Drawing on qualitative data from clinicians in Australia and the UK, the study moves beyond event-based views of discharge to illuminate the distributed, often invisible practices through which discharge safety is constructed in everyday clinical care, and to identify the organisational conditions that support or constrain it.

## 3. The Study

This study aims to conceptualise discharge readiness in CCUs as anticipatory nursing and interprofessional work and to examine how professional roles, organisational systems and service contexts shape discharge across two national settings.

## 4. Method

### Design

This study employed a qualitative, exploratory design to examine health-care professionals’ perspectives on discharge practices in coronary care units (CCUs) in Australia and the United Kingdom (UK). The study was guided by an interpretivist epistemology and a contextualist ontology, recognising that discharge practices are actively formed from professional, organisational and system-level contexts. Data were analysed using reflexive thematic analysis (RTA) (Braun & Clarke, 2006, 2019, 2022). Reporting was informed by the COREQ checklist (Tong et al., 2007) in line with journal requirements, and by the Reflexive Thematic Analysis Reporting Guidelines (RTARG; Braun & Clarke, 2024) to support methodological coherence (see Supplementary File 1).

### Study Setting, Participants and Inclusion Criteria

Participants were recruited from coronary care units in Australia and the UK. The two settings were selected to enable interpretive contrast across organisationally distinct but structurally comparable cardiac care contexts. In Australia, recruitment took place across one site in Western Australia, comprising a tertiary referral hospital unit serving mixed urban and regional populations, with 20 beds. This unit was considered broadly representative of Coronary Care Unit (CCU) services and had established clinical-academic relationships. In the UK, recruitment took place across two CCU site(s) in North-West England, comprising tertiary referral CCU serving metropolitan populations, including a dedicated 10 bed CCU and 26 coronary care beds within an acute hospital setting.

Purposeful sampling was used to support variation in professional background, gender and years of experience. Clinicians were eligible if they had at least three years of CCU experience and were actively involved in patient care and discharge-related decision-making. In Australia, eligible professional groups included registered nurses, senior charge nurses, allied health professionals, medical staff and pharmacists, reflecting the multidisciplinary composition of CCU teams in the participating units. In the UK, recruitment was restricted to nursing staff, reflecting both the central coordinating role of nurses in CCU discharge in the UK context and the feasibility constraints of data collection during the post-acute phase of the COVID-19 pandemic (2021-2022). The resulting difference in sample composition across the two settings was treated as a design feature of interpretive contrast rather than an equivalence failure, with the intent of illuminating shared underlying processes and how local organisational scaffolding shapes their expression.

Thirty-seven clinicians participated overall. In Australia, 24 staff contributed through seven focus groups and two individual interviews (one medical, one allied health), comprising 14 nursing staff, 6 allied health professionals and 4 medical staff. Nineteen (79%) were female. The mean age was 37.5 years (range 28–57), with mean professional experience of 12.5 years (range 4–37); most worked full-time. In the UK, 13 nurses (7 male, 6 female) participated in individual interviews; all worked full-time, with a mean age of 38.7 years (range 28–50) and mean professional experience of 13.2 years (range 5–25). Detailed demographic characteristics are presented in Table 1.

**Table 1.**
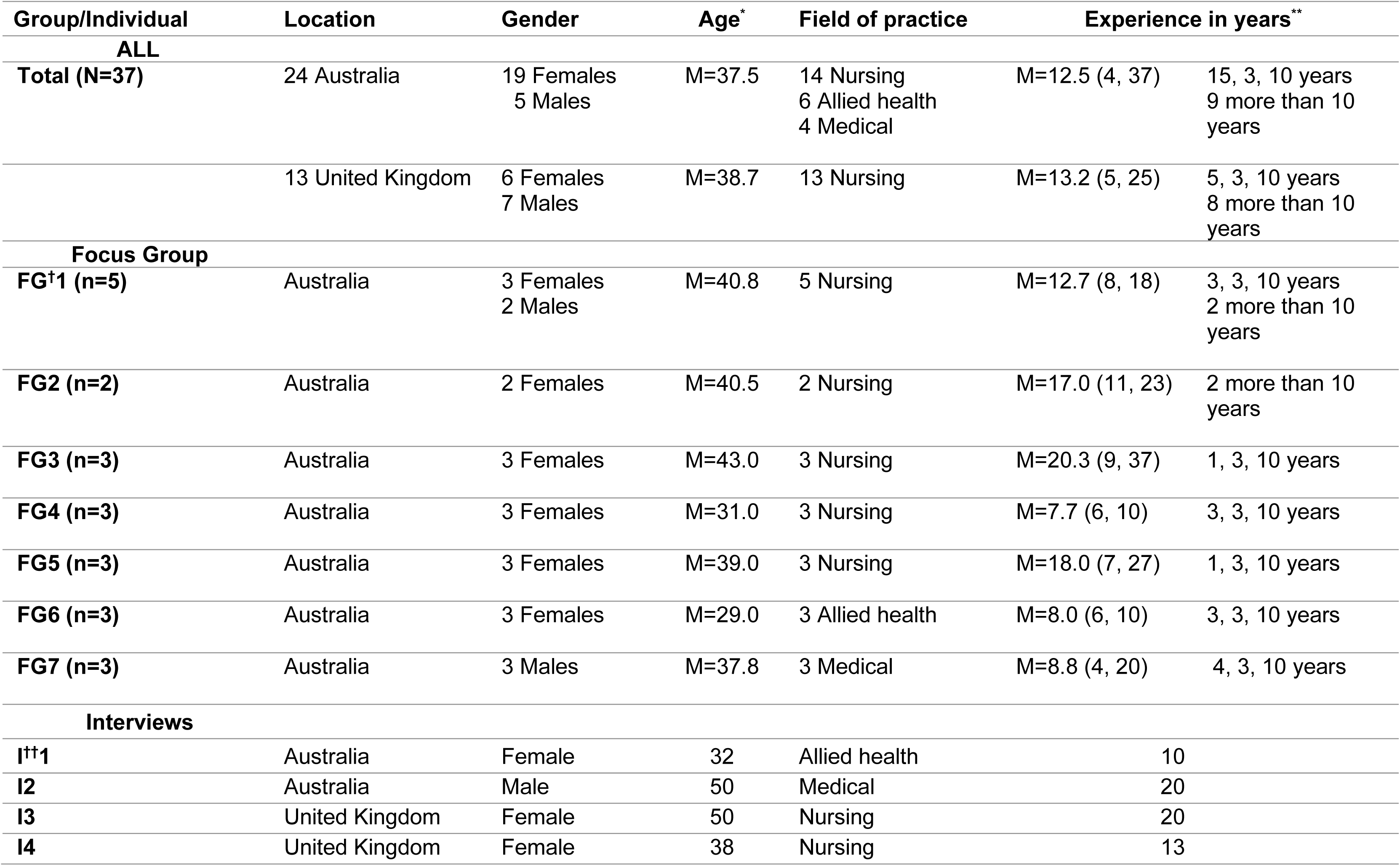
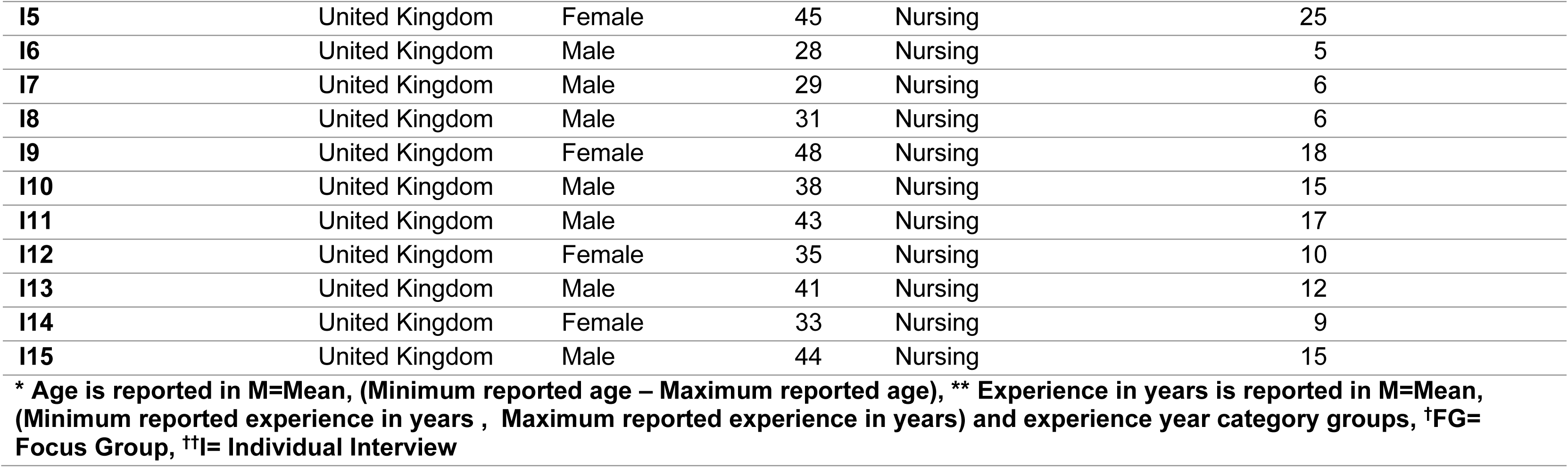
Demographic characteristics of participants (n=37)

| Group/Individual | Location | Gender | Age* | Field of practice | Experience in years** |  |
| --- | --- | --- | --- | --- | --- | --- |
| ALL |  |  |  |  |  |  |
| Total (N=37) | 24 Australia | 19 Females<br>5 Males | M=37.5 | 14 Nursing<br>6 Allied health<br>4 Medical | M=12.5 (4, 37) | 15, 3, 10 years<br>9 more than 10 years |
|  | 13 United Kingdom | 6 Females<br>7 Males | M=38.7 | 13 Nursing | M=13.2 (5, 25) | 5, 3, 10 years<br>8 more than 10 years |
| Focus Group |  |  |  |  |  |  |
| FG†1 (n=5) | Australia | 3 Females<br>2 Males | M=40.8 | 5 Nursing | M=12.7 (8, 18) | 3, 3, 10 years<br>2 more than 10 years |
| FG2 (n=2) | Australia | 2 Females | M=40.5 | 2 Nursing | M=17.0 (11, 23) | 2 more than 10 years |
| FG3 (n=3) | Australia | 3 Females | M=43.0 | 3 Nursing | M=20.3 (9, 37) | 1, 3, 10 years |
| FG4 (n=3) | Australia | 3 Females | M=31.0 | 3 Nursing | M=7.7 (6, 10) | 3, 3, 10 years |
| FG5 (n=3) | Australia | 3 Females | M=39.0 | 3 Nursing | M=18.0 (7, 27) | 1, 3, 10 years |
| FG6 (n=3) | Australia | 3 Females | M=29.0 | 3 Allied health | M=8.0 (6, 10) | 3, 3, 10 years |
| FG7 (n=3) | Australia | 3 Males | M=37.8 | 3 Medical | M=8.8 (4, 20) | 4, 3, 10 years |
| Interviews |  |  |  |  |  |  |
| I††1 | Australia | Female | 32 | Allied health | 10 |  |
| I2 | Australia | Male | 50 | Medical | 20 |  |
| I3 | United Kingdom | Female | 50 | Nursing | 20 |  |
| I4 | United Kingdom | Female | 38 | Nursing | 13 |  |

|  |  |  |  |  |  |
| --- | --- | --- | --- | --- | --- |
| **I5** | United Kingdom | Female | 45 | Nursing | 25 |
| **I6** | United Kingdom | Male | 28 | Nursing | 5 |
| **I7** | United Kingdom | Male | 29 | Nursing | 6 |
| **I8** | United Kingdom | Male | 31 | Nursing | 6 |
| **I9** | United Kingdom | Female | 48 | Nursing | 18 |
| **I10** | United Kingdom | Male | 38 | Nursing | 15 |
| **I11** | United Kingdom | Male | 43 | Nursing | 17 |
| **I12** | United Kingdom | Female | 35 | Nursing | 10 |
| **I13** | United Kingdom | Male | 41 | Nursing | 12 |
| **I14** | United Kingdom | Female | 33 | Nursing | 9 |
| **I15** | United Kingdom | Male | 44 | Nursing | 15 |

**\* Age is reported in M=Mean, (Minimum reported age – Maximum reported age), \*\* Experience in years is reported in M=Mean, (Minimum reported experience in years , Maximum reported experience in years) and experience year category groups, †FG= Focus Group, ††I= Individual Interview**

### Recruitment

Recruitment followed local governance arrangements in each country. In Australia, the Nurse Unit Manager and Clinical Nurse Consultant distributed study information via email and displayed posters in the CCUs. In the UK, a member of the research team circulated the information pack to all eligible staff. Potential participants received a study information sheet and consent form and were invited to contact the research team directly. Interviews and focus groups were scheduled at times compatible with clinical duties and conducted during work hours with organisational approval.

### Data collection

Data were collected between July 2021 and December 2022 by X in Australia or X in the UK. Both researchers hold nursing or health-related backgrounds relevant to the study context. Semi-structured interviews and focus groups were conducted either face-to-face in private hospital rooms or via secure videoconferencing (Microsoft Teams). Sessions lasted between 45 and 90 minutes. A mixed approach was adopted to maximise participation across two systems facing differing operational pressures. Focus groups allowed exploration of shared professional norms and interprofessional dynamics, while individual interviews enabled more personalised accounts of organisational constraints.

The interview guide (see Supplementary File 2) was developed collaboratively during a research workshop and refined with input from an advisory group of experienced CCU clinicians. It covered discharge practices, factors influencing discharge decisions, characteristics of high-quality discharge planning, and opportunities to improve transitional care. Three pilot interviews refined question wording and ordering and were retained in the final dataset. Field notes captured contextual observations and supported reflexive analysis. Although the broader study also explored readmission, the present paper focuses exclusively on discharge processes and coded separately to maintain analytic focus.

### Data analysis

Transcripts were produced verbatim, anonymised and managed using NVivo 12 (QSR International, 2018). Analysis was conducted using reflexive thematic analysis (RTA) (Braun & Clarke, 2006, 2019, 2022) and proceeded iteratively in stages. Transcripts from each country were read repeatedly for familiarisation, with initial coding led by X and conducted separately for each dataset to preserve contextual specificity and analytic independence. Early coding decisions were discussed regularly with XX to surface interpretive assumptions, before codes were clustered into candidate themes supported by memo writing and team discussions examining coherence and conceptual boundaries. Themes were then developed into interpretative accounts for each dataset, followed by conceptual refinement undertaken collaboratively by X and XXX, interrogating relationships between themes, clarifying boundaries, and strengthening theoretical links.

A subsequent cross-site interpretive integration was performed by X and XXX. Rather than merging or re-coding the datasets into a single framework, the two analyses were compared to identify patterns of similarity and difference in how clinicians described discharge work, developing higher-level interpretations while maintaining the epistemological integrity of RTA and allowing organisational and system-level influences to become visible through interpretive contrast (Braun & Clarke, 2022). This approach reflects qualitative scholarship on interpretive integration across related datasets (Sandelowski & Barroso, 2007; Thorne, 2016). (See Supplementary File 3 for extended quotations and interpretive commentary).

The research team spanned nursing practice, health psychology, allied health and clinical medicine across both national contexts. Primary analysis was led by X, whose nursing background-oriented attention towards the organisational context of discharge work and the practical realities of everyday clinical practice. XXX grounding in health psychology brought complementary attention to the emotional, relational and cognitive dimensions of anticipatory clinical work and to how professional identity shapes clinical action. Interpretive assumptions were surfaced and examined through team discussion, memo writing and reflexive engagement across the broader team, and we recognise that alternative interpretations remain possible.

### Ethical considerations

Ethical approval was granted by the Edith Cowan University Human Research Ethics Committee, Western Australia (reference number 2020-02076) and South Metropolitan Health Service Research Ethics Committee, Western Australia (reference number RGS0000000 3676). Participants received full study information, were assured of confidentiality and their right to withdrawal, and provided written or emailed consent. Verbal consent was also reconfirmed at the beginning of each session. All transcripts were de-identified and stored securely on password-protected systems accessible only to the research team.

### Findings

The four themes presented below were generated from analysis of interviews and focus groups with the 37 clinicians described in the Methods (see Table 1). The themes depict discharge as a complex organisational, relational and clinical process enacted across professional boundaries (anticipatory, ongoing labour shaped by system demands, documentation structures and interdisciplinary negotiation). Illustrative extracts from both national contexts are included throughout, with attention to where the underlying mechanisms were shared and where their expression diverged.

### Theme 1: Discharge as Anticipatory Nursing Work

Across both countries, nurses depicted discharge preparation embedded into everyday nursing work, characterised by persistent monitoring, administrative organisation and coordination with patients, families and other services. Rather than describing discharge as a single endpoint decision, participants framed readiness as something that had to be actively maintained and continuously worked towards across the admission trajectory. While this orientation towards readiness was shared across the UK and Australia, its expression varied between systems.

In Australia, nurses emphasised the use of structured tools, particularly bedside checklists and discharge booklets, to scaffold preparation throughout the patient journey. These artefacts enabled tracking of functional progress, belongings, educational components and home readiness indicators. Discharge work was normalised as methodical, procedural and built into routine documentation. The checklist served not only as a record but as a forward-looking tool that prompted attention to issues that might affect discharge before they became urgent:

> “What’s really important when you are looking at discharges is that we as nurses are really proactive from day one… In every bed, we have a checklist so they can be ready for discharge at any time.” (AUS, FG1 Nurse)

This structured approach contributed to perception among nurses that they held primary responsibility for ensuring all aspects of discharge preparation had been completed. Several participants described performing an implicit “auditing” role to guarantee that gaps in readiness were identified early:

> “I can just feel like we’re the only ones who make sure everything’s done before they go home. We double check, going through all the list to confirm everything.” (AUS, FG1 Nurse)

In contrast, UK nurses rarely referenced formal checklist systems. Instead, they described discharge as an ongoing process of monitoring, escalation and coordination that began at admission and continued throughout the patient stay:

> “And the way I look at discharge, as soon as someone’s admitted you are starting the discharge process from the day that they come in.” (UK Nurse 4)

UK participants emphasised the importance of identifying potential barriers early and coordinating interdisciplinary input to support safe planning:

> “Early assessment of individual patients gives us a chance to identify their functional needs to determine appropriate support services when they return home such as coordination with interdisciplinary team physiotherapist, rehabilitation services, counselling”. (UK Nurse 8)

Nurses also described proactively monitoring discharge trajectories and identifying concerns or opportunities relating to discharge preparation:

> “Sometimes we’ll flag early discharges with the doctors.” (UK Nurse 9)

Their accounts highlighted ongoing judgement work in anticipating functional, logistical and social issues that might disrupt discharge readiness if left unresolved.

Despite these differences, there was overlap across settings. Across both, participants described substantial judgement work in balancing competing priorities, managing time pressures and responding to patients’ preparedness, motivation and understanding. Participants in both countries also described the cognitive, emotional and professional demands of holding and sustaining this work overtime. One Australian nurse described carrying significant emotional burden while simultaneously coordinating care, filling gaps and ensuring continuity (AUS, Nurse, FG2). Others described responsibility for addressing logistical and social barriers extending beyond narrowly clinical tasks:

> “It’s pretty much for us to work out problems and then ask to fix them… how are they getting home… driving restrictions.” (UK Nurse 11)

Discharge readiness was experienced not as a discrete clinical assessment but as sustained anticipatory labour, coordinative, problem-solving and relational in nature, maintained across the full admission and carrying a cognitive and emotional burden evident across both settings.

### Theme 2: Discharge as Collective Accomplishment

Discharge was rarely described as a single clinical decision made by one professional group. Instead, participants across both countries portrayed discharge readiness as something negotiated through ongoing interaction between nurses, doctors, pharmacists, allied health professionals and patients, with different forms of expertise contributing to assessment of whether discharge was safe and feasible. Readiness was therefore experienced as a collective accomplishment produced through coordination across the multidisciplinary team.

Nurses portrayed themselves as central to initiating and sustaining these discussions. Their continuous contact with patients afforded them a longitudinal understanding of functional progression, symptom stability, social circumstances and psychosocial needs, which they routinely brought into discharge planning conversations. UK nurses, in particular, emphasised the need to prompt medical review when they perceived patients to be approaching readiness for discharge:

> "You often have to flag with the doctors… we’ve done this, this and this, and this patient is certainly suitable for discharge." (UK Nurse 6)

This reflects how nurses’ ongoing monitoring informed negotiations about readiness, positioning them as active contributors to discharge decision-making, not as passive implementers of medical plans. Producing alignment across professional groups was itself described as a form of work, often initiated and coordinated by nursing staff.

Participants also described how influence within these negotiations could vary according to professional hierarchy and clinical experience. Some nurses described feeling that their contributions carried greater weight when they were more senior or well established within the team:

> "Quite a lot… you have influence depending on your level of experience." (UK Nurse 1)

Others described variability in how collaborative discharge discussions were enacted in practice, with some medical staff actively seeking nurse perspectives while others communicated discharge decisions more hierarchically:

> "Sometimes you’ve got doctors who actually seek your opinion… other times it’s just an order." (UK Nurse 15)

These accounts suggest that discharge collaboration was a negotiated process influenced by interpersonal relationships, professional authority and organisational culture. Allied health professionals were positioned as important contributors to discharge assessment, particularly in relation to functional readiness and rehabilitation needs. UK nurses highlighted the value of early physiotherapy and occupational therapy involvement in supporting coordinated and safe discharge planning:

"If physio and OT get involved from the beginning, any issues can be discussed with the team, so together we make an informed discharge decision." (UK Nurse 1)

Pharmacists in the Australian sample were positioned as particularly influential in determining whether discharge was practically and clinically safe, especially in relation to medication access, adherence and continuity of treatment following discharge. Their assessments extended beyond prescription review to include practical considerations such as weekend access to medication supply, patient adherence and service availability:

> "If I have no concerns, I’ll say they’re good to go. But if they can’t get medications on the weekend or they’re non-compliant… then I’ll say, hold on, that’s not appropriate." (AUS Nurse, FG2)

Medical staff also acknowledged the importance of pharmacy input within the discharge planning process:

> "The ability of the pharmacist to review each patient and explain the medications is very important before discharge." (AUS Doctor, FG7)

While medical participants recognised the contributions of other disciplines, their accounts tended to frame discharge in terms of clinical authorisation and safety oversight rather than as a negotiated and coordinated process. This distinction is analytically meaningful. It suggests that professional role shapes not only what clinicians contribute to discharge but how the work itself is understood, with nurses and allied health staff inhabiting a coordinative logic and medical staff a gatekeeping one. Where these orientations operated in alignment, they supported effective discharge; where they diverged, the coordinative labour of nursing staff became less visible and less legible to those with formal discharge authority.

Despite the emphasis on collaborative working, participants across both settings described communication gaps and coordination failures that could delay discharge or create uncertainty around readiness. These accounts highlight how discharge decisions depended not only on clinical judgement but also on the effectiveness of information sharing and team coordination:

> "Another issue is inconsistencies in communicating among health care providers leading to further delay and increase unnecessary burden on our workload, then poor coordinator in our role within interdisciplinary team can create prolong delay in discharge process." (AUS Nurse, FG2)

Although this extract is drawn from the Australian dataset, similar concerns about fragmented communication and unclear responsibility were evident in the UK accounts. Participants described discharge readiness as vulnerable to delays when interdisciplinary communication broke down or when responsibilities for coordination were insufficiently defined.

Overall, discharge readiness was co-constructed through distributed clinical expertise and ongoing interdisciplinary negotiation. Nursing input frequently anchored these processes through continuous patient oversight and coordination work, while pharmacists and allied health professionals shaped critical dimensions of medication safety, functional readiness and practical feasibility. Although the underlying processes were broadly shared across settings, differences in emphasis were evident: UK participants highlighted early functional assessment and interdisciplinary input, whereas Australian accounts foregrounded medication management and logistical constraints, particularly around access to medications.

### Theme 3: Discharge Documentation as a Site of Tension

If discharge readiness was co-constructed through interprofessional negotiation, documentation functioned as one of the primary mechanisms through which that coordination was recorded, communicated and operationalised. Participants across both countries described documentation as a significant component of discharge practice, viewed as both essential to continuity of care and a source of considerable demand on clinicians’ time. This dual role positioned documentation as a site of ongoing tension, shaping workflow, time allocation and communication quality.

Australian clinicians, in particular, expressed frustration with the volume and perceived complexity of documentation required. Lengthy discharge summaries were often perceived as poorly aligned with the practical communication needs of patients and community providers, while also competing with time for direct patient interaction:

> “I don’t understand why we need to write ten-page discharge summaries… that’s too much information. Instead of focusing on identifying patients’ needs and ensuring they understand their follow-up care.” (AUS Doctor, FG7)

This critique reflected a perceived disconnect between procedural expectations and clinicians’ understanding of meaningful discharge communication. Rather than viewing documentation simply as informational transfer, participants frequently contrasted administrative completion with relational aspects of discharge preparation, including patient education, reassurance and personalised explanation. Nurses similarly framed documentation demands as displacing time they would otherwise dedicate to patient education and reassurance:

> “Constantly working on paperwork and coordination of these paperwork causes further delays in the discharge process, instead of this, we could spend more time to educate our patients one by one and provide consistent care for our patients”. (AUS, Nurse, FG1)

Several UK participants framed documentation in terms of how it competed with relational and coordinative work that they saw as more central to discharge:

> “Discharge summaries… should be a very succinct, clear document that tells anyone exactly what they need in a few lines.” (UK Nurse 9)

This emphasis on brevity reflects a pragmatic orientation towards communication. In contrast to Australian accounts, UK participants appeared less likely to strongly foreground documentation as a source of excessive workload, suggesting subtle contextual differences in how documentation was experienced and articulated across the settings. Across both contexts, participants viewed documentation as central to discharge communication yet poorly aligned with the realities of time pressures and workflow demands. For some clinicians, documentation requirements were experienced as adding layers of administrative work that extended beyond what they considered clinically meaningful discharge activity:

> “Completing lots of documents resulted in significant delays in discharge process, this documentation only adds an extra administrative layer to our job on top of what we do”. (AUS Doctor, FG7)

Documentation requirements were often experienced as externally imposed rather than locally negotiable, even where their fit with patient and community needs was openly questioned. This points to a deeper tension running through these accounts: two competing logics operating simultaneously within discharge work. An administrative logic frames documentation as institutional compliance, the auditable, visible trace of a process completed to system requirements. A clinical logic frames it as communication, a means of ensuring that what the team knows travels with the patient beyond the ward. Patient education, reassurance, personalised explanation, become displaced and this is consequential for discharge readiness but leaves no equivalent institutional trace. Documentation is therefore not simply an informational tool but an organisational mechanism that determines what discharge work is seen to consist of and, implicitly, what remains invisible within it.

### Theme 4: Discharge Within System and Organisational Pressures

Participants repeatedly situated discharge within wider system and organisational pressures, describing practices shaped as much by these structural conditions as by clinical readiness. Across both countries, clinicians reported working in environments characterised by high patient turnover, limited bed capacity and fluctuating staffing levels, all of which exerted pressure to expedite discharge.

Australian participants emphasised practical constraints related to medication access, rural geography and reduced weekend service provision. These factors shaped what was considered a “safe” discharge and, at times, delayed patient departure despite clinical stability:

> “On weekends in the regional or rural areas… there is a significant barrier to accessing pharmacy services including waiting for medications, lack of service hours, [and] even pharmacist staff. We should be relied on better management for transport, staffs and medications to operate successfully”. (AUS Nurse, FG2)

Participants also described how organisational capacity varied across the week, creating periods where discharge processes become more difficult to coordinate:

> “Weekend services are very different… we’re much shorter staffed.” (UK Nurse 10)

These constraints were not incidental inconveniences but expressions of the system architecture within which Australian CCUs operate: a federated, geographically dispersed healthcare structure in which the practical conditions for safe discharge, medication supply, service access, and transport, could not always be assumed to be in place once the patient left the unit. For Australian clinicians, anticipatory discharge work therefore included planning around what the system might not be able to provide, rather than simply what the patient needed clinically.

UK clinicians described pressures associated with patient throughput, bed demand and limitations in community support services. Delays were frequently linked to difficulties securing timely follow-up care, community nursing or social care provision for patients who were otherwise clinically stable:

> “Delays in discharge process could be because of connecting patients to relevant services in the community setting, especially those who need an extra support in the community”. (UK Nurse 10)

Several participants described situations where organisational pressures surrounding bed availability influenced discharge decision-making itself:

> “Maybe about … half… of our discharges, comes down to bed pressure.” (UK Nurse 4)

These accounts suggest that discharge timing was not determined solely by clinical criteria but was also through wider institutional pressures to maintain patient flow and organisational capacity. Discharge readiness is not simply a clinical state to be reached and confirmed but a negotiated position within a system whose imperatives do not always align with individual patient need, and which clinicians must navigate, not simply deliver.

> “If someone is ready Friday afternoon they may stay until Monday.” (UK Nurse 5)

The UK accounts reveal a different configuration of the same underlying constraint.

Where Australian clinicians navigated access gaps at the periphery of the system, UK clinicians described pressure emanating from the centre, from the institutional imperative to maintain bed flow in a system where community and social care capacity frequently could not keep pace with clinical demand. Discharge readiness was bounded not by geography or weekend pharmacy provision, but by whether the downstream system had the capacity to receive a patient who was otherwise ready to leave. The explicit naming of bed pressure as a driver of discharge timing suggests that operational necessity had become a co-determinant of readiness alongside clinical criteria. Across both settings, discharge was framed as enacted within system and organisational conditions that clinicians could anticipate but not themselves resolve. The nature of the constraint differed, access and practical feasibility in Australia, downstream capacity and institutional throughput in the UK, but the underlying mechanism was shared: the rate-limiting step on discharge is something the CCU itself cannot fix. What varies between systems is where that limit falls and therefore what anticipatory work must address. Participants described tension between facilitating safe, well-supported transitions and meeting operational demands that limited the time available for discharge preparation. These accounts suggest that discharge safety depends not only on clinical readiness but on the alignment between patient state and system capacity, an alignment that clinicians could anticipate and work towards but could not themselves produce. Where system-level conditions did not align with patient need, no amount of anticipatory or interprofessional work could substitute for absent services. This finding reframes discharge readiness as fundamentally contingent: not a property of the patient that clinicians assess, but a relational achievement produced through the fit between what the patient needs and what the surrounding system can provide at that moment. The implication is significant for how discharge work is understood and resourced. Anticipatory nursing labour and interprofessional coordination are necessary conditions for safe discharge but are not, by themselves, sufficient ones.

### Comparative Summary Across Themes

The four themes were developed within each national dataset before being brought into interpretive dialogue through cross-site integration. Although the datasets differed in professional composition and data collection format, this staged process was not designed to rank or directly compare two equivalent samples, but to use contrast as an analytic resource. Where divergence was observed, it illuminated how the same underlying processes take different forms depending on local organisational scaffolding. Where convergence was observed, it suggested that anticipatory work, interprofessional negotiation, documentation tension and system constraint reflect something more fundamental about how discharge readiness is produced in high-acuity, short-stay contexts, regardless of national setting. The cross-national design therefore does not describe two different discharge systems so much as reveal a shared set of mechanisms whose surface expression varies with context.

## 6. Discussion

Discharge from coronary care units in Australia and the UK was described not as a final event but as a continuous, anticipatory process woven throughout the admission, coordinated, negotiated and revised in real time as clinical and organisational conditions shifted. Nurses held this work together, maintaining an overarching view of clinical status, functional recovery and psychosocial needs while linking the contributions of medical, pharmacy and allied health teams. By illuminating this in-admission organisational labour, the study extends transitional care research and brings Allen’s (2014) account of invisible nursing work into the high-acuity, short-stay context of coronary care.

### Discharge as anticipatory work

These findings challenge the dominant transitional care models and mechanistic accounts of care transitions (Coleman et al., 2006; Naylor et al., 2011; Zwart et al., 2021), that treat discharge mainly as a handover event or intervention, by showing how readiness is built up across the admission rather than enacted at its end. The analysis aligns closely with Allen’s (2014) account of the invisible organisational work that nurses perform to hold complex care together, and with Strauss and colleagues’ (1985) framing of articulation work and invisible work (Star and Strauss 1999), describing the unseen activity of fitting together the contributions of different actors into a workable whole. The preparation in everyday nursing routines, operating as unremarkable practice than as deliberate procedure, is consistent with what May and Finch (2009) describe within Normalisation Process Theory as collective action.

The distinctive contribution here is the setting. Short admissions, high acuity and dense interdependence between disciplines compress preparation into a constantly renegotiated process that is unusually visible. Coronary care therefore offers a revealing site for examining how nursing coordination interacts with organisational and system pressure.

Australian nurses often relied on bedside checklists and discharge booklets to scaffold preparation. UK nurses emphasised earlier, iterative engagement with allied health professionals. The tools and routines differed, but the underlying orientation of keeping discharge in view and acting ahead of it remained consistent. Recognising discharge as ongoing preparation rather than a final sign-off has clear practice implications. When only sign-off is resourced, the upstream work that makes safe discharge possible stays invisible and under-supported. Participants’ accounts that nursing staff were "the only ones who make sure everything’s done" suggest that, in current practice, this work is held but not often acknowledged.

A health psychology lens sharpens what anticipatory discharge work involves at the level of the individual clinician. Psychologically, the orientation nurses described, holding discharge readiness in mind across an entire admission, tracking evolving indicators of patient preparedness, and acting on anticipated need before problems arise, is a form of prospective planning: cognitively demanding, future-directed and sustained across competing immediate demands. Maintaining this orientation in the high-acuity, rapid-turnover environment of coronary care involves not only clinical expertise but active cognitive self-regulation, particularly where workforce pressures reduce the time available for unhurried assessment. Cognitive load of this kind is distributed across the team and the organisational artefacts that structure anticipatory work in both settings.

Understood in this light, the tension participants described around documentation is not simply about time: it is about the displacement of cognitive scaffolding that supports anticipatory work by administrative demands that record rather than enable it. Participants’ accounts of emotional burden, sustained vigilance and felt responsibility for ensuring nothing was missed align with the literature on emotional labour in nursing, the management of feeling as an integral component of professional role performance (Theodosius, 2008; Smith, 2012). Participants did not describe this emotional and cognitive load as exceptional. It was experienced as an ordinary feature of the role, suggesting it had become normalised alongside the anticipatory work itself, with implications for workforce wellbeing that warrant attention in future research.

### Discharge as collective accomplishment

No single profession owned the discharge decision. Readiness was arrived at through interaction across nursing, medical, pharmacy and allied health roles, each contributing a partial and discipline-specific perspective on whether discharge was safe and feasible. These perspectives were aligned through informal conversation and negotiation more often than formal protocols. These accounts describe collective sensemaking in Weick’s (1995) terms: readiness was not a fixed clinical state retrieved through individual assessment, but a judgement constructed and continuously revised through interprofessional exchange. Where that exchange broke down, through communication gaps, unclear responsibility or hierarchical asymmetry, the shared understanding of readiness faltered with it. The data add specificity to existing accounts of interprofessional coordination in acute care and to qualitative evidence on what facilitates or constrains care transitions in practice (Allen, 2014; Scott et al., 2017). In Australia, pharmacists were positioned as gatekeepers of medication safety, with explicit authority to delay discharge when supply, adherence or weekend access raised concerns. UK accounts foregrounded allied health professionals, particularly physiotherapy and occupational therapy, as upstream contributors whose early engagement shaped discharge timing. These patterns are best read as illustrations of how role configurations get embedded in routine discharge work, not as defining features of either system. The shared understanding of readiness produced through interprofessional negotiation still had to travel beyond the original conversation, across shifts and to community providers and professional groups not present. Documentation was the primary mechanism through which it did, and it carried its own demands and tensions.

### Documentation as organisational mechanism

Documentation was both essential to continuity of care and a recurring source of tension. Australian clinicians described lengthy summaries that consumed time better spent on patient education and felt poorly aligned to what community providers needed. UK participants raised concerns about clarity and usability. In both countries, documentation functioned as more than information transfer: it shaped how time was allocated, what counted as completion and how discharge work was prioritised against competing demands. This is the sociotechnical dimension of discharge, the way administrative artefacts and system infrastructure shape clinical practice and resonates with Allen (2014) who describes the burden that record systems impose on nursing organisational labour. That this burden was experienced across both national settings, despite differences in the tools and workflows involved, underlines its structural rather than incidental character.

### System and organisational conditions

Beyond the documentation system, broader structural pressures shaped what was achievable in any given case. Both samples described environments of high turnover, limited beds and fluctuating staffing. Australian teams additionally navigated weekend pharmacy access gaps and the particular demands of rural distances; UK teams struggled with securing community nursing and social care for patients who were stable but could not be safely discharged without downstream support in place (Glasby et al., 2021; Shepperd et al., 2013). Organisational and system conditions determined how much anticipatory work was needed to produce a safe discharge in any given case, independently of clinical status.

These cross-national contrasts are best read not as a ranking of two systems but as an illustration of how anticipatory discharge work, a shared underlying process, is influenced by local organisational scaffolding.

### Conceptual representation of the findings

The themes describe a process that is both distributed and layered, shaped simultaneously by individual clinical judgement, interprofessional negotiation, administrative infrastructure and system context. What follows draws these elements into an integrated interpretive account of how discharge readiness is enacted in coronary care.

The findings are represented as a data-grounded conceptual model of discharge readiness in coronary care (Figure 1). Conceptual models developed from qualitative data serve as interpretive frameworks for integrating and communicating patterns across themes. The model here is therefore offered in this tradition rather than as a formal or generalisable theory (Maxwell, 2013). The model positions anticipatory nursing work as the foundational layer of discharge labour, with interprofessional negotiation and documentation operating alongside it, each intensifying as discharge approaches. System and organisational conditions form the surrounding context shaping what is achievable at any point across the admission. The visual representation is chosen because it conveys something the written themes cannot easily express in sequence: the simultaneous operation of multiple processes across a shared admission trajectory, each with a distinct temporal character. The nested structure is deliberate, conveying that system conditions do not simply coexist with clinical work but determine its outer limits. The model therefore adds to the text not by introducing new claims but by rendering the relationships between the four themes legible as an integrated whole. It’s central claim, and the study’s principal contribution to transitional care theory, is that underlying processes are shared across both settings while their expression varies with local organisational scaffolding.

**Figure 1:**
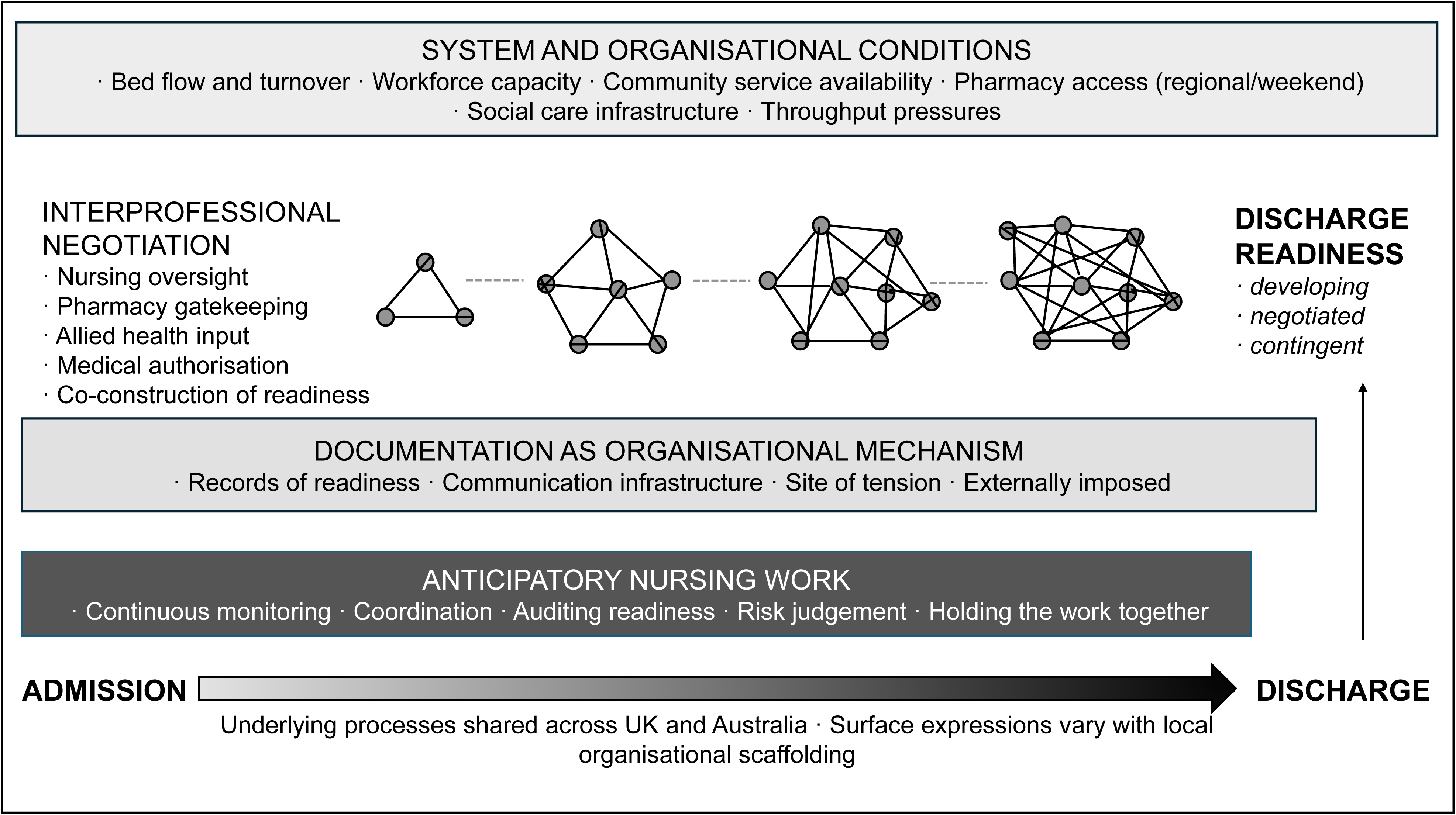
Discharge readiness in coronary care: anticipatory work across the admission within system and organisational conditions. The figure maps the four themes derived through clinician accounts in CCUs in Australia and the United Kingdom: anticipatory nursing work (Theme 1) is depicted as continuous across the admission; interprofessional negotiation (Theme 2) and documentation as organisational mechanism (Theme 3) operating alongside this labour and intensifying as discharge approaches; and system and organisational conditions (Theme 4) forming the contextual layer within which all clinical activity is situated.

### Implications for practice and policy

Across both settings, the findings suggest that improving discharge in CCU contexts requires attention to the conditions through which anticipatory work is produced, not only the procedures through which it is recorded. Protected time for early multidisciplinary input, reliable access to pharmacy and allied health, and streamlined documentation are likely to be more consequential than further refinement of the discharge moment itself. Explicit recognition of nurses as coordinators is equally important; without it, the labour that makes safe discharge possible remains invisible and under-resourced. Brief discharge-focused multidisciplinary check-ins offer one practical mechanism for supporting the alignment work the findings identified.

Within the UK context, participants’ emphasis on early functional assessment and interdisciplinary input reflects national guidance on transitions of care and hospital discharge (NICE, 2015; Department of Health and Social Care, 2022) and reinforces the role of the established allied health and community nursing infrastructure. The persistent constraints around community service capacity reflect wider structural pressures on social care documented elsewhere (Glasby et al., 2021), and which set practical limits on what clinical effort alone can achieve. Streamlining discharge documentation to prioritise clarity and clinical relevance is consistent with longstanding evidence on effective information transfer (Kripalani et al., 2007) and directly addresses the documentation tensions this study identified.

Within the Australian context, participants’ accounts of documentation burden align with ACSQHC standards advocating clear and patient-centred discharge communication (ACSQHC, 2017, 2020). The system-level constraints around medication access, particularly in regional and weekend contexts, point to pharmacy infrastructure as a structural determinant of discharge feasibility rather than a downstream administrative concern. Workforce planning that recognises and explicitly resources the coordination embedded in nursing discharge work would directly address the labour identified in the findings (Naylor et al., 2011).

Internationally, the findings support policy approaches that conceptualise discharge as a coordinated longitudinal process rather than a discrete event, consistent with the WHO Framework on Integrated People-Centred Health Services (WHO, 2016). Improvement strategies are unlikely to transfer cleanly between systems without attention to the local organisational conditions through which discharge work is enacted. A clinician-facing checklist developed directly from the findings (Supplementary File 4) is offered as an illustrative, adaptable practice tool rather than a validated intervention.

### Strengths and limitations

Strengths include the interdisciplinary research team, inclusion of multiple professions in the Australian sample, and reflexive thematic analysis with careful within- and cross-site interpretive integration. These perspectives supported analytic depth, with nursing expertise grounding the analysis in clinical realities and health psychology extending its interpretive reach into behavioural and organisational dimensions. Reflexive and collaborative analysis, including discussion of interpretive assumptions and analytic decision-making, enhanced coherence and rigour. The dual-country design sharpened insight into organisational influences on discharge and supports the potential transferability of findings to comparable high-acuity settings. The study offers an empirically grounded conceptualisation of discharge readiness, and a practice-oriented checklist derived directly from the analysis.

Several limitations should be acknowledged. The two datasets were not symmetrical: the UK sample comprised nurses only, interviewed individually, while the Australian sample included multiple professions and was conducted mostly through focus groups. This reflected local feasibility constraints and a deliberate design response to context rather than an equivalence failure and observed contrasts between settings are interpreted cautiously accordingly. The study captures clinician perspectives only; future research examining the experiences of patients and family is warranted. Sites were specific CCUs and may not represent all cardiac settings. Finally, in keeping with reflexive thematic analysis, the themes are interpretive constructs, and alternative interpretations are possible. The conceptual model and associated checklist remain preliminary, requiring further testing of feasibility and implementation in practice.

## 7. Conclusion

Discharge in coronary care is better understood as continuous anticipatory work than as an endpoint. Across two healthcare systems, clinicians described preparation that began at admission and was sustained through ongoing assessment, coordination, negotiation and risk judgement, with nurses holding much of the connective work. The cross-national contrast exposed the influence of local organisational scaffolding without revealing fundamentally different processes. Strengthening discharge in CCU contexts is therefore likely to depend less on refining the discharge moment than on supporting the staffing, access, and documentation conditions through which readiness is produced.

Testing the conceptual model in settings beyond coronary care would establish how far it travels and where the underlying account needs revision. Clinician accounts alone cannot establish whether the preparation described here translates into readiness that patients and families actually experience. Evaluating the checklist in practice would shift attention from whether the practices it describes are sound to what conditions allow them to be enacted.

## Data Availability

All data produced in the present study are available upon reasonable request to the authors.

## Acknowledgements

We thank the health professionals who generously contributed their time, insights and experience to this study. Their openness in discussing the complexities of discharge practice made this research possible. We also acknowledge the support of the participating hospitals and departmental teams who facilitated recruitment and provided access to clinical settings. Finally, we are grateful to colleagues who offered methodological and conceptual guidance during the development of the study.

## Supplementary Files

1. **COREQ and RTARG**
2. **Interview Guide**
3. **Data Extracts and Analytical Development**
4. **Discharge Checklist**

## Supplementary File 1

### COREQ–RTARG Integrated Checklist

**Reporting frameworks:** COREQ (Tong et al., 2007) and Reflexive Thematic Analysis Reporting Guidelines (RTARG; Braun & Clarke, 2024).

This integrated checklist was developed to satisfy journal requirements for COREQ reporting whilst remaining methodologically consistent with Reflexive Thematic Analysis Reporting Guidelines (RTARG; Braun & Clarke, 2024). Where COREQ items are not reported, explanatory commentary is provided to indicate whether omission reflects the epistemological and methodological commitments of reflexive thematic analysis rather than incomplete reporting. The checklist should therefore be interpreted as a mapping exercise between COREQ and RTARG rather than a conventional COREQ compliance checklist.

### Status categories

- Reported
- Reported with RTARG adaptation
- Not reported – justified through RTARG
- Not applicable

| Topic | Item No. |  | Guide Question / Description | Section Heading in Manuscript | Status | RTARG Alignment / Commentary |
| --- | --- | --- | --- | --- | --- | --- |
| **Domain 1: Research Team and Reflexivity** |  |  |  |  |  |  |
| Personal characteristics | 1 |  | Which author(s) conducted the interviews / focus groups? | Methodology – Data Collection | **Reported** | Data collection was led by AR (Amineh Rashidi), who conducted the Australian focus groups and individual interviews. UK individual interviews were conducted by [initials to confirm]. All sessions were conducted by members of the wider research team with relevant clinical or academic backgrounds. Consistent with COREQ and RTARG transparency expectations. |
|  | 2 |  | What were the researcher's credentials? | Methodology – Data Analysis; Author Contributions | **Reported** | Interdisciplinary team spanning nursing practice, health psychology, allied health and clinical medicine across both national contexts. AR's nursing background and LN's health psychology grounding explicitly acknowledged as shaping analytic orientation. Consistent with both COREQ and RTARG. |
|  | 3 |  | Occupation at time of study? | Methodology – Data Analysis; Author | **Reported** | Professional roles noted in the Methods. AR identified as lead researcher with a nursing background; LN as a health psychologist. Full team credentials outlined in author |
|  |  |  |  | Contributions |  | contributions. Consistent with COREQ. |
|  | **4** |  | Researcher gender? | Research Team and Reflexivity | **Not reported – justified through RTARG** | Not reported intentionally. Consistent with RTARG, reflexive thematic analysis prioritises researcher positioning, assumptions and analytic engagement over demographic description alone. Positionality is therefore addressed through reflexive accounts of disciplinary, professional and theoretical positioning rather than demographic characteristics. |
|  | **5** |  | Experience and training? | Research Team and Reflexivity; Methodology – Data Analysis | **Reported with RTARG adaptation** | Team expertise described in broad terms. AR's nursing background and LN's health psychology grounding are explicitly noted as shaping analytic attention and interpretive reach respectively. RTARG aligns with reflexive practice rather than credentialism. |
|  | **6** |  | Relationship established prior to study? | Methodology – Study Setting and Participants; Recruitment | **Reported** | Established clinical-academic relationship at the Australian site facilitated recruitment via the Nurse Unit Manager and Clinical Nurse Consultant. In the UK, a research team member circulated the information pack. Reflexivity regarding these access arrangements is acknowledged in the analytic sections, consistent with RTARG. |
| Relationship with participants | **7** |  | What did participants know about the researcher? | Methodology – Data Collection; Ethical Considerations | **Reported** | Participants received full study information sheets detailing the purpose of the research, team background and data use prior to agreeing to participate. Verbal consent reconfirmed at the start of each session. Consistent with COREQ. |
|  | **8** |  | Interviewer characteristics – assumptions, assumptions, biases? | Research Team and Reflexivity; Methodology-Data Analysis | **Reported** | Reflexivity statement embedded in the Methods (Data Analysis section). AR's nursing orientation explicitly noted as directing analytic attention toward organisational context; LN's health psychology perspective noted as extending interpretive reach into cognitive and emotional dimensions of anticipatory work. Consistent with RTARG's emphasis on owning and making productive use of researcher perspectives. |
| **Domain 2: Study Design** |  |  |  |  |  |  |
| Theoretic<br>al<br>framewor<br>k | **9** |  | Methodologi<br>cal<br>orientation<br>and theory<br>described? | Method<br>ology-Design | **Reported** | Reflexive Thematic Analysis (RTA; Braun & Clarke, 2006, 2019, 2022) within an interpretivist epistemology and contextualist ontology, recognising that discharge practices are actively shaped by professional, organisational and system-level contexts. Explicitly aligned with RTARG expectations and Braun & Clarke's own framing of RTA. |
| Participan<br>t selection | **10** |  | How were participants selected? | Method<br>ology –<br>Study<br>Setting<br>and<br>Particip<br>ants | **Reported** | Purposive sampling to support variation in professional background, gender and years of experience. Eligibility criteria: minimum three years of CCU experience and active involvement in discharge-related decision-making. Consistent with COREQ and RTARG. |
|  | **11** |  | How were participants approached ? | Method<br>ology –<br>Recruit<br>ment | **Reported** | Australia: Nurse Unit Manager and Clinical Nurse Consultant distributed study information by email and poster display. UK: a research team member circulated the information pack to all eligible staff. Participants contacted the research team directly to express interest. Consistent with COREQ. |
|  | **12** |  | Sample size? | Method<br>ology –<br>Study<br>Setting<br>and<br>Particip<br>ants;<br>Table 1 | **Reported** | 37 clinicians overall: 24 in Australia (7 focus groups and 2 individual interviews; 14 nurses, 6 allied health, 4 medical staff) and 13 nurses in the UK (individual interviews). Sample size adequacy addressed with reference to Braun & Clarke's guidance on data adequacy and richness. Detailed characteristics in Table 1. |
|  | **13** |  | Non-participation – numbers declining? | Method<br>ology –<br>Recruit<br>ment | **Reported** | No refusals reported. All participants who received the information pack and made contact with the research team consented to participate. RTARG emphasis on data adequacy rather than numerical target is consistent with the reporting. |
| Setting | **14** |  | Where were data collected? | Method<br>ology –<br>Study<br>Setting<br>and<br>Particip | **Reported** | Coronary care units within tertiary referral hospitals. Australia: one site in Western Australia (20-bed CCU). UK: two sites in North-West England (10-bed dedicated CCU and 26 |
|  |  |  |  | ants;<br>Data<br>Collecti<br>on |  | coronary care beds within an acute hospital). Sessions conducted face-to-face in private hospital rooms or via Microsoft Teams videoconferencing. |
|  | **15** |  | Presence of non-participants during data collection? | Method<br>ology –<br>Data<br>Collecti<br>on | **Partial.**<br>**Reported** | UK individual interviews: interviewer and participant only. Australian focus groups: multiple participants and facilitator present by design; this is inherent to the focus group method and supports exploration of shared professional norms and interprofessional dynamics. Consistent with RTARG. |
|  | **16** |  | Sample characteristics? | Table 1;<br>Method<br>ology –<br>Study<br>Setting<br>and<br>Particip<br>ants | **Reported** | Detailed demographic characteristics reported in Table 1: age, gender, field of practice, years of experience, employment status, and data collection format for all 37 participants. Professional roles and CCU settings described in the Methods section. |
| **Domain 2: Data Collection** |  |  |  |  |  |  |
| Data collection | **17** |  | Interview guide used and provided? | Supple<br>mentary<br>File 2 –<br>Interview<br>and<br>Focus<br>Group<br>Schedule | **Reported** | Semi-structured interview and focus group schedule provided as Supplementary File 2. Covers discharge practices, factors influencing discharge decisions, high-quality discharge planning and transitional care improvement. Developed collaboratively at a research workshop, refined with input from an advisory group of experienced CCU clinicians, and piloted (three pilot interviews retained in the final dataset). Consistent with COREQ and RTARG. |
|  | **18** |  | Repeat interviews conducted? | Method<br>ology –<br>Data<br>Collecti<br>on | **Not**<br>**applicabl**<br>**e** | Single-session data collection only. Consistent with RTARG's position that data adequacy and richness, rather than repeated participant contact, are the appropriate criteria in RTA. |
|  | **19** |  | Audio or visual recording? | Method<br>ology –<br>Data<br>Collecti<br>on;<br>Ethical<br>Consid<br>erations | **Reported** | Sessions audio-recorded using encrypted devices or secure videoconference recording. Transcripts produced verbatim, anonymised and managed in NVivo 12. Consistent with COREQ. |
|  | **20** |  | Field notes made? | Method<br>ology –<br>Data<br>Collecti | **Reported** | Field notes captured contextual observations during data collection. Reflexive memos recorded throughout analysis to |
|  |  |  |  | on;<br>Data<br>Analysi<br>s |  | surface and examine interpretive assumptions. Both are central to the epistemological commitments of RTA and to RTARG. |
|  | **21** |  | Duration of interviews / focus groups? | Method<br>ology-Data<br>Collecti<br>on | **Reported** | Sessions lasted between 45 and 90 minutes. A mixed approach was adopted – focus groups in Australia and individual interviews in the UK – to maximise participation across two systems facing differing operational pressures. |
|  | **22** |  | Data saturation discussed? | Method<br>ology-Study<br>Setting<br>and<br>Particip<br>ants;<br>Data<br>Analysi<br>s | **Reported with RTARG adaptation** | Addressed as data adequacy in line with Braun & Clarke (2021/2022). Adequacy framed in terms of richness and depth of data generated rather than numerical saturation. Consistent with RTARG and current RTA scholarship. |
|  | **23** |  | Transcripts returned to participants for correction? | Method<br>ology-Data<br>Analysi<br>s | **Not reported-justified through RTARG** | Transcripts were not returned for participant checking, consistent with RTARG, which does not recommend member checking as a validity strategy in RTA. Credibility of the data generation process was supported through involvement of an advisory group of experienced CCU clinicians in the development of the interview schedule. Analytical rigour maintained through team discussion, memo writing and reflexive engagement. |
| **Domain 3: Analysis and Findings** |  |  |  |  |  |  |
| Data analysis | **24** |  | How many coders?<br>Description of coding process? | Method<br>ology-Data<br>Analysi<br>s | **Reported with RTARG adaptation** | Initial coding led by AR, with early coding decisions regularly discussed with MD (Melissa Dunham) to surface interpretive assumptions. Candidate themes developed through memo writing and team discussion. Cross-site interpretive integration performed by AR and LN. RTARG discourages inter-coder reliability as a validity metric in RTA; collaborative discussion and reflexive engagement are the appropriate mechanisms. |
|  | **25** |  | Coding tree / theme development provided? | Supple<br>mentary<br>File 3-Data<br>Extract<br>s and<br>Analytic | **Reported** | Supplementary File 3 provides data extracts with initial analytic observations, interpretive commentary for each theme and a cross-theme integration table mapping mechanisms to the conceptual model. Supports |
|  |  |  |  | al<br>Develo<br>pment |  | analytic transparency consistent with RTARG. |
|  | **26** |  | Themes identified in advance or derived from data? | Method<br>ology -<br>Data<br>Analysi<br>s | **Reported** | Inductive theme generation through iterative coding and interpretive engagement with the data. No a priori themes imposed. Consistent with the epistemological commitments of RTA and RTARG expectations. |
|  | **27** |  | Software used? | Method<br>ology -<br>Data<br>Analysi<br>s | **Reported** | NVivo 12 (QSR International, 2018) used for transcript management and coding. Mind maps and team discussions also used to visualise analytic patterns and support theme development. RTARG expresses no preference regarding software. |
|  | **28** |  | Participant feedback on findings? | Method<br>ology -<br>Data<br>Analysi<br>s | **Not**<br>**reported**<br>**-justified**<br>**through**<br>**RTARG** | No participant feedback on findings was sought, consistent with RTARG, which does not advocate member checking as a validity strategy in RTA. The credibility of the data generation process was supported through involvement of an advisory group of experienced CCU clinicians in developing the interview schedule. Analytical rigour maintained through team discussion, reflexive memo writing and collaborative interpretive integration. |
|  | **29** |  | Participant quotations used to illustrate themes? | Finding<br>s;<br>Supple<br>mentar<br>y File 3 | **Reported** | Extensive direct quotations included throughout the Findings, attributed by participant identifier. Supplementary File 3 provides additional extracts with analytic commentary for each theme. Consistent with COREQ and RTARG expectations for evidential transparency. |
|  | **30** |  | Is data and findings consistent? | Finding<br>s;<br>Discuss<br>ion | **Reported** | Coherence between data extracts and interpretive accounts demonstrated throughout the Findings. Cross-site integration maintained the epistemological integrity of RTA whilst using interpretive contrast as an analytic resource to illuminate shared mechanisms and surface-level variation between settings. |
|  | **31** |  | Major themes clearly and fully described? | Finding<br>s | **Reported** | Four themes presented as full interpretive accounts: (1) Discharge as Anticipatory Nursing Work; (2) Discharge as Collective Accomplishment; (3) |
|  |  |  |  |  |  | Discharge Documentation as a Site of Tension; (4) Discharge Within System and Organisational Pressures. Each supported by data extracts, analytic commentary and a cross-theme integration table in Supplementary File 3. Consistent with RTARG quality guidance. |
|  | **32** |  | Minor themes or negative cases described? | Finding s; Supple mentar y File 3 | **Reported with RTARG adaptation** | Cross-national variation in how shared underlying processes were expressed is described and interpreted as illuminating variation rather than treated as negative cases in the traditional sense. Examples include pharmacist gatekeeping in Australia versus allied health prominence in the UK, and rural geographic constraints versus downstream throughput pressure. Supplementary File 3 provides additional analytic detail. Consistent with Braun & Clarke's quality guidance as reflected in RTARG. |

## Supplementary File 2

### Interview and Focus Group Schedule

Exploring Perceptions and Practices around Coronary Care Unit (CCU)

### Opening

- Welcome and thank you for your participation.
- Introduce the purpose: *“This study seeks to explore experiences, perceptions, and strategies regarding patient readmissions to the Coronary Care Unit (CCU). We are interested in understanding what contributes to discharge and readmissions processes*
- Explain confidentiality, the voluntary nature of participation, audio-recording, and data handling.
- Confirm **verbal consent** before proceeding.

### Section A: Demographic Information

#### To help contextualise the findings, we would like to collect some background information. (in focus group confirm individually)

1. Gender:
2. Age:
3. Years of experience in healthcare:
4. Current employment status (e.g. full-time, part-time, agency):
5. Years in your current role or position:
6. Highest level of education:

### Section B: Discharge Practices

7. *How are decisions about discharging a patient from the CCU typically made? (*Prompts: Which team members are involved? What criteria guide readiness for discharge?
8. *What challenges do you encounter when planning or coordinating discharge?* Prompts: Patient understanding, family readiness, community services, communication barriers.
9. How is information typically communicated to patients at discharge? Prompts: How do you check understanding (e.g., medication changes, symptoms, follow-up)?/ What challenges arise when providing discharge information?
10. How do you determine whether a patient feels ready and able to manage after discharge? Prompts: Health literacy cues; Confidence in medication management
11. Understanding of red-flag symptoms; Family or carer support
12. What organisational or systemic issues affect the discharge process? Prompts: Staffing levels, Time pressures, Bed pressures or early discharge, Inconsistent communication across shifts, Access to community services
13. How well do you feel discharge processes link with primary care or community services? Prompts: GP communication Community nursing Cardiac rehabilitation referrals
14. How do you involve family members or informal supports in discharge planning? Prompts: Inclusion in discussions Assessing carer readiness, When support is limited or absent
15. *How does the discharge process affect the likelihood of readmission?*

Prompts: Continuity of care, follow-up, medication issues, health literacy.

16. *What would help improve discharge safety and reduce gaps in transitional care?*
17. What changes or support would help you deliver safer and more effective discharge processes?”
18. Have you seen any approaches or innovations elsewhere that support better discharge and reduce unplanned readmissions?

### Section C: Understanding of Readmissions

This section explores participants’ general perceptions of readmission and their relevance in the CCU.

19. In general, what are your thoughts on readmission?
20. Prompt: What does this mean to you? Could you please tell me what you see as the main reasons for patients being readmitted to the CCU? Prompt: Are there other reasons why you think patients are readmitted?
21. Tell me about a time a patient you cared for was readmitted. Prompt: How did that situation unfold and how was it managed?
22. Have you noticed a common pattern for readmissions? *Prompts: Why do you think that happens? Could you give an example from your own experience?*

### Section D: Current Practice and Influencing Factors (20–25 minutes)

23. What kind of problems do you see with discharge processes and also readmissions to CCU? *Prompt: This could relate to patients, families, staffing, care processes, or system issues*.
24. Can you describe how decisions are typically made about discharge/ readmissions? *Prompts: Who is involved in those decisions? What role do different professionals play?*
25. What factors tend to influence whether a patient is readmitted? *Prompts: Consider patient needs, family/carer circumstances, system capacity, discharge planning*.
26. How are readmissions currently managed in your team? Prompts: Are there any formal or informal processes for review?

### Section E: Opportunities for Improvement

27. How do you think that the readmission rate can be reduced in the CCU? *Prompt: What interventions or support do you think might help?(at discharge?)*
28. If you could change or improve one thing to prevent unnecessary readmissions, what would it be?/ to improve discharge?
29. What support would help you or your team in preventing readmissions?
30. Have you seen or heard of effective strategies elsewhere that could work here?

### Wrap-Up

- Do you have any final reflections or points you’d like to add?
- Thank participant(s) again, offer information for follow-up if needed, and end the session.

### Supplementary File 3: Data Extracts and Analytical Development

This supplementary file presents illustrative quotations and analytic commentary for each of the four themes generated through reflexive thematic analysis (Braun & Clarke, 2006, 2019, 2022). For each theme, a table maps data extracts to initial analytic observations and their interpretative contribution to the emerging theme. Analytic commentary following each table describes how coding decisions developed into interpretative accounts. The file concludes with a cross-theme integration table summarising the mechanisms identified and each theme’s contribution to the conceptual model of discharge readiness.

The extracts are drawn from focus groups and individual interviews with 37 clinicians across coronary care units in Australia (n = 24) and the United Kingdom (n = 13). Participant identifiers follow the conventions used in the main manuscript: AUS participants are identified by professional group and focus group number (e.g. AUS, FG1 Nurse; AUS Pharmacist, FG2); UK participants are identified by sequential number (e.g. UK Nurse 4). The supplementary file is intended to support analytic transparency and to illustrate the interpretive movement from data to theme consistent with reflexive thematic analysis.

### Theme 1: Discharge as Anticipatory Nursing Work

Across both countries, nurses framed discharge preparation as an ongoing process embedded in everyday clinical work, characterised by persistent monitoring, practical coordination and anticipatory problem-solving from admission through to discharge. The following extracts illustrate this orientation and the range of activities through which it was enacted.

| Data Extract | Initial Analytic Observation | Interpretative Contribution to Theme |
| --- | --- | --- |
| *"What's really important when you are looking at discharges is that we as nurses are really proactive from day one... In every bed, we have a checklist so they can be ready for discharge at any time."* (AUS, FG1 Nurse) | Structured tool-based anticipatory approach embedded in Australian CCU routines | Illustrates how bedside checklists scaffold proactive discharge preparation from admission, treating readiness as an ongoing property of each patient rather than a terminal assessment |
| *"I can just feel like we're the only ones who make sure everything's done before they go home. We double check, going through all the list to confirm everything."* (AUS, FG1 Nurse) | Implicit auditing role assumed by nursing staff | Demonstrates nurses' perceived primary responsibility for ensuring discharge completeness, with a felt sense of ownership that extended beyond formally allocated tasks |
| *"And the way I look at discharge, as soon as someone's admitted you are starting the discharge process from the day that they come in."* (UK Nurse 4) | Discharge framed as beginning at the point of admission rather than at clinical sign-off | Demonstrates the anticipatory orientation underpinning discharge work, readiness understood as a process built across the admission rather than confirmed at its end |
| *"Early assessment of individual patients gives us a chance to identify their functional needs to* | Proactive identification of functional and social needs from admission | Illustrates how anticipatory work extends beyond clinical stability to |
| *determine appropriate support services when they return home such as coordination with interdisciplinary team physiotherapist, rehabilitation services, counselling.*” (UK Nurse 8) |  | encompass service coordination and post-discharge support, beginning early in the patient journey |
| *“Sometimes we’ll flag early discharges with the doctors.”* (UK Nurse 9) | Nurses proactively initiating discharge trajectories with medical staff | Demonstrates nurses operating as initiators of the discharge process rather than passive recipients of medical decisions, a form of forward-directed interprofessional communication |
| *“It’s pretty much for us to work out problems and then ask to fix them... how are they getting home... driving restrictions.”* (UK Nurse 11) | Nurses coordinating practical and logistical discharge barriers | Reveals hidden coordination work within everyday nursing practice: practical problem-solving that falls outside formal clinical task descriptions but is essential to safe discharge |
| *“Sometimes you’ll get told they’re going home... they’re going home now... in half an hour.”* (AUS CCU Nurse) | Sudden and unpredictable timing of discharge decisions despite anticipatory preparation | Highlights core tension between proactive nursing preparation and the reactivity imposed by rapid or late discharge decisions, anticipatory work disrupted by organisational timing |
| *“You’re often chasing your tail to find the answers.”* (AUS CCU Nurse) | Workload intensity and cognitive complexity of coordination | Reveals the practical and cognitive labour underpinning discharge readiness, coordination experienced as effortful and reactive despite anticipatory intent, generating visible workload burden |

### Analytic Commentary

Initial coding captured recurrent references to *planning ahead*, *anticipating discharge needs* and *monitoring readiness*. Iterative engagement with the dataset revealed that clinicians consistently described discharge preparation as beginning at the point of admission rather than at the point of clinical sign-off. Nurses maintained awareness of patient progress across the full admission trajectory, anticipating potential obstacles, coordinating referrals and addressing practical barriers before they became consequential.

These activities were interpreted as a form of anticipatory nursing labour that underpins discharge readiness across the admission. Importantly, participants simultaneously described situations in which discharge decisions were imposed suddenly, due to clinical deterioration, senior medical review or organisational bed pressure, creating tension between proactive preparation and reactive decision-making. This juxtaposition between anticipation and unpredictability was interpreted as a core feature of discharge work rather than an incidental variation.

The theme integrates two interrelated dimensions: anticipatory preparation enacted by nurses across the admission, and the unpredictable timing of formal discharge decisions within complex care systems. Across both settings, this labour was characterised by cognitive and emotional demands that had become normalised as ordinary features of the nursing role.

### Theme 2: Discharge as Collective Accomplishment

Discharge readiness was rarely described as the sole determination of any one professional group. Instead, participants depicted it as negotiated through ongoing interaction across nursing, medical, pharmacy and allied health roles, each contributing distinct expertise to the collective judgement of whether discharge was safe and feasible.

| Data Extract | Initial Analytic Observation | Interpretative Contribution to Theme |
| --- | --- | --- |
| *"You often have to flag with the doctors... we've done this, this and this, and this patient is certainly suitable for discharge."* (UK Nurse 6) | Nurses prompting and shaping medical discharge decision-making | Illustrates nurses' active role in discharge decisions, positioning them as contributors and advocates rather than passive implementers of medical plans |
| *"Quite a lot... you have influence depending on your level of experience."* (UK Nurse 1) | Professional seniority mediating influence within interprofessional negotiations | Highlights how hierarchy and experience shape whose voice carries weight in discharge discussions, a structural feature of negotiated readiness that affects equity of contribution |
| *"Sometimes you've got doctors who actually seek your opinion... other times it's just an order."* (UK Nurse 15) | Variability in collaborative practice shaped by individual clinician relationships | Demonstrates that interprofessional negotiation was context-dependent, shaped by individual relationships and unit culture rather than by fixed structural protocols |
| *"If physio and OT get involved from the beginning, any issues can be discussed with the team, so together we make an informed discharge decision."* (UK Nurse 1) | Early allied health involvement enabling collective and informed discharge assessment | Demonstrates that discharge readiness was understood as a distributed assessment requiring contributions across multiple disciplines, with timing of involvement shaping the quality of decisions |
| *"If I have no concerns, I'll say they're good to go. But if they can't get medications on the weekend or they're non-compliant... then I'll say, hold on,"* | Pharmacists acting as gatekeepers of medication safety and practical feasibility | Shows how pharmacy expertise shaped discharge timing beyond clinical prescription review, incorporating |
| *that's not appropriate.*" (AUS Pharmacist, FG2) |  | access, adherence and system-level supply concerns, particularly salient in regional and weekend contexts |
| *"The ability of the pharmacist to review each patient and explain the medications is very important before discharge."* (AUS Doctor, FG7) | Medical recognition of pharmacy's contributory role in discharge authorisation | Illustrates interprofessional acknowledgement of specialist contributions to collective discharge readiness, framed within medical oversight, suggesting awareness of distributed expertise even where authority remained hierarchical |
| *"Another issue is inconsistencies in communicating among health care providers leading to further delay and increase unnecessary burden on our workload, then poor coordinator in our role within interdisciplinary team can create prolong delay in discharge process."* (AUS Nurse, FG2) | Communication gaps and coordination failures generating delay and additional burden | Demonstrates that co-construction of readiness was vulnerable to breakdown, coordination failure a source of discharge delay and compounded workload, underscoring the fragility of interprofessional alignment |

### Analytic Commentary

Early coding identified patterns relating to *communication between professional groups*, *influence within discharge decisions* and *collaborative care planning*. Through interpretative analysis, these were integrated into the concept of discharge readiness as a collective accomplishment, something produced through ongoing dialogue rather than confirmed by any single professional assessment.

Nurses were analytically positioned as central to initiating and sustaining interprofessional discharge discussions, drawing on their continuous contact with patients to bring a longitudinal perspective into negotiations. The data also revealed that professional hierarchies shaped whose voice carried most weight: some clinicians described actively seeking nursing input; others communicated discharge decisions as directives. This variability was interpreted as reflecting negotiated rather than structurally fixed collaboration. Pharmacists (in the Australian dataset) and allied health professionals (particularly in the UK dataset) shaped distinct dimensions of readiness assessment, medication safety and practical feasibility in the former; functional readiness and rehabilitation needs in the latter. These role-specific contributions illustrated how collective readiness was built from distributed expertise, whilst the accounts of communication failure and unclear responsibility illustrated the fragility of this co-construction when interprofessional alignment broke down.

### Theme 3: Discharge Documentation as a Site of Tension

Documentation was described by participants across both countries as essential to continuity of care but also as a source of considerable demand on clinicians’ time and a recurring source of friction within discharge work. The following extracts illustrate the tension between administrative and clinical logics that documentation brought into view.

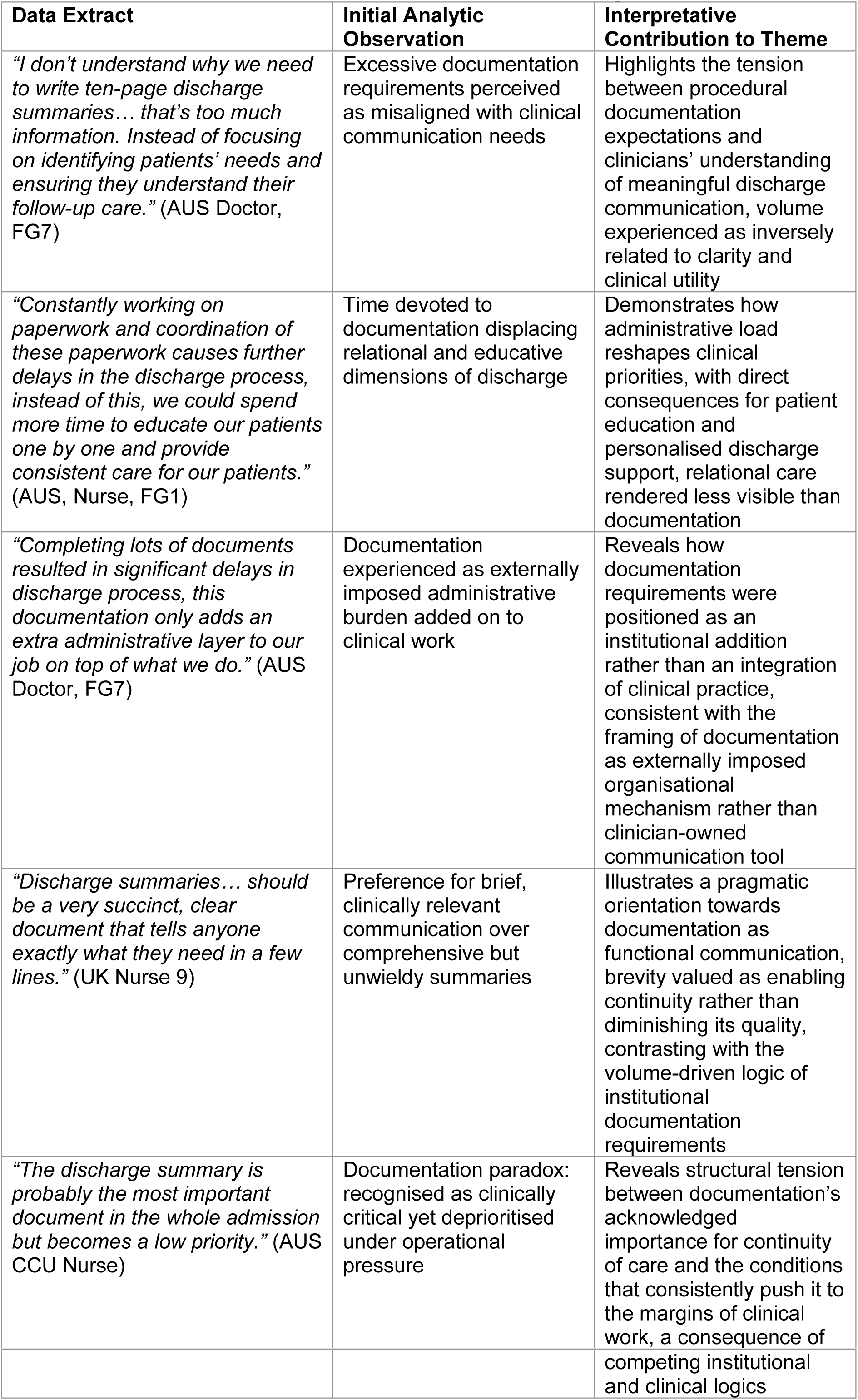

### Analytic Commentary

Initial coding captured references to *paperwork burden*, *documentation workload* and *information transfer*. As analysis developed, participants’ accounts revealed a consistent ambivalence: documentation was simultaneously recognised as the primary mechanism through which discharge information travelled beyond the ward and experienced as poorly aligned with the practical communication needs of patients and receiving providers. The interpretative framing of documentation as a site of organisational tension emerged from this dual character. Two competing logics operated simultaneously: an administrative logic, which framed documentation as institutional compliance and the auditable trace of a completed process; and a clinical logic, which framed it as communication, a means of ensuring that what the team understood about a patient travelled with them into the community. When administrative demands dominated, relational and educative dimensions of discharge preparation were displaced, leaving no equivalent institutional trace. Documentation requirements were experienced by participants as externally imposed and difficult to challenge locally, even where their alignment with patient and community need was openly questioned. This positions documentation not merely as an informational tool but as an organisational mechanism that structures what discharge work consists of what is made visible, and by implication, what remains invisible within it.

### Theme 4: Discharge Within System and Organisational Pressures

Participants situated discharge work within wider system and organisational conditions that shaped what was achievable in any given case. Both Australian and UK accounts described structural constraints on discharge feasibility that operated independently of clinical readiness and that no amount of anticipatory or interprofessional work could themselves resolve.

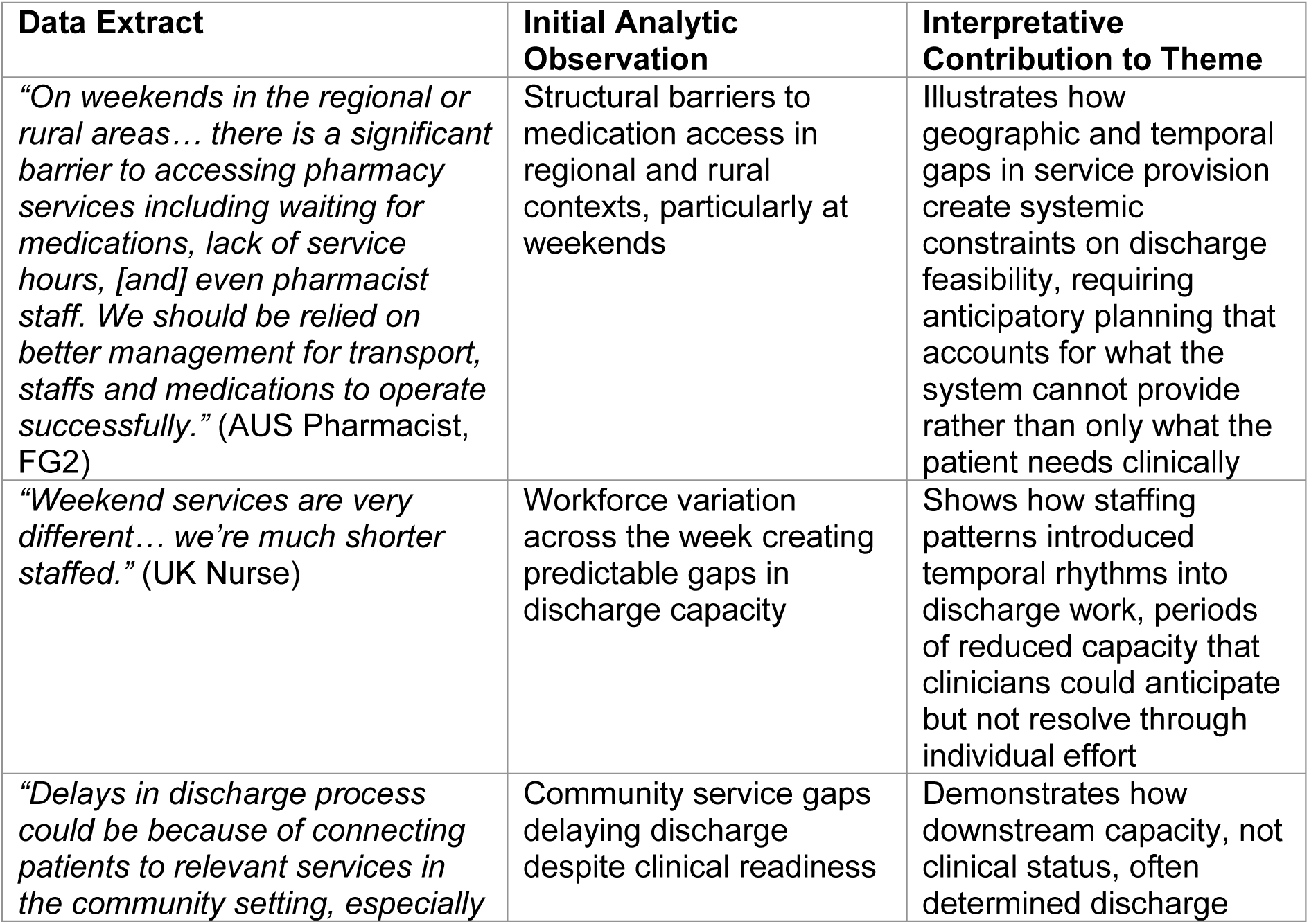
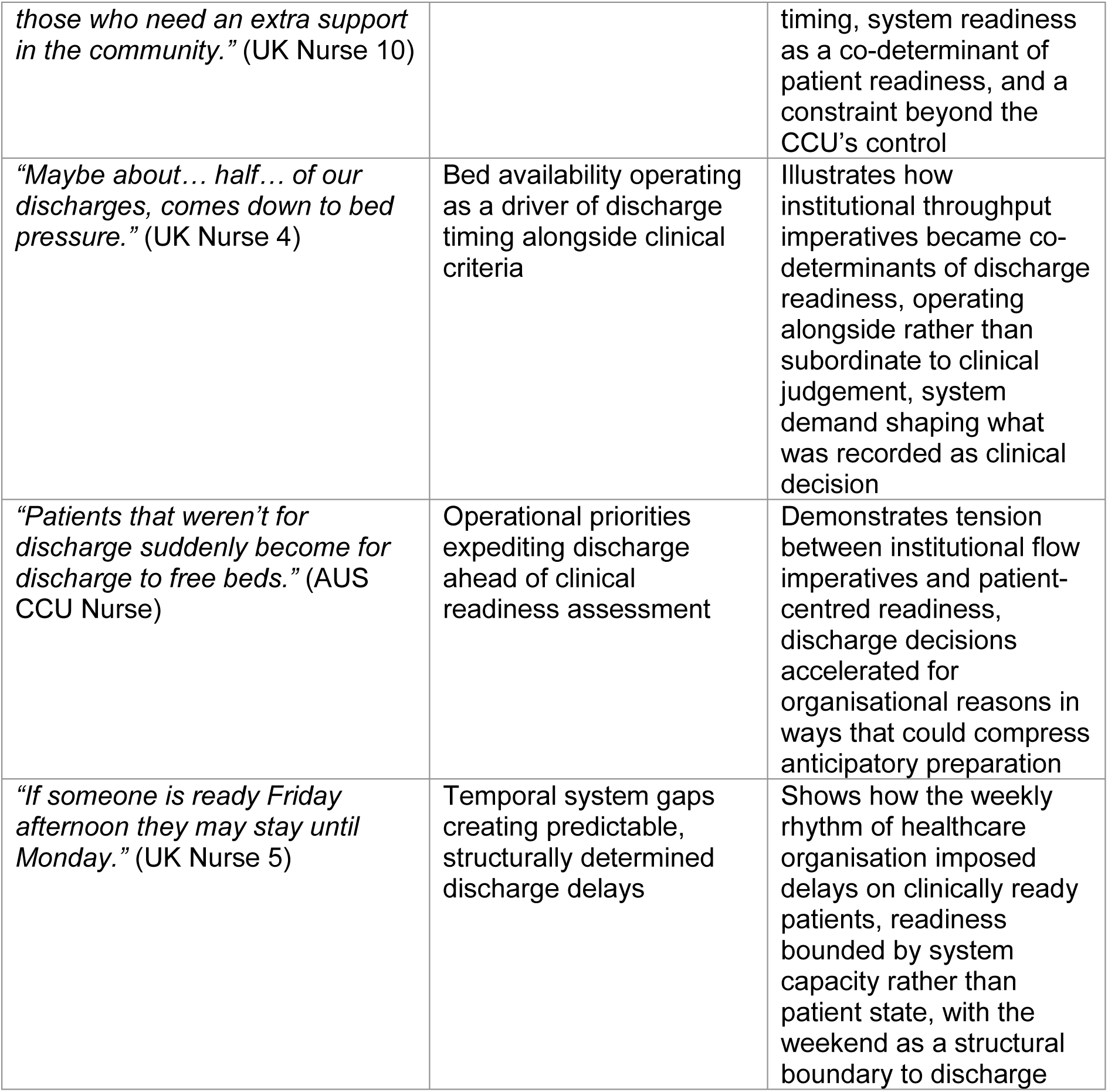

### Analytic Commentary

Coding initially identified repeated references to *bed pressures*, *service availability*, *staffing levels* and *resource limitations*. These were interpreted as reflecting broader organisational logics shaping discharge practice, conditions through which discharge work was enacted rather than incidental inconveniences.

Across both settings, clinicians described navigating competing priorities between patient safety and system throughput. The nature of the structural constraint differed between datasets: Australian clinicians navigated access and practical feasibility gaps at the periphery of a geographically dispersed system (pharmacy supply, transport, rural distance); UK clinicians described throughput pressure and community capacity failures emanating from the institutional centre. Despite these different configurations, the underlying mechanism was shared: the rate-limiting step on discharge was frequently beyond the CCU’s own control.

This finding reframes discharge readiness as fundamentally contingent, not a property of the patient that clinicians assess and confirm, but a relational achievement produced through the alignment between what the patient needs and what the surrounding system can provide at that moment. Anticipatory nursing labour and interprofessional coordination are necessary conditions for safe discharge but not, by themselves, sufficient ones.

### Cross-Theme Integration: Contribution to the Conceptual Model

The table below summarises the key empirical insight, the mechanism identified and each theme’s contribution to the conceptual model of discharge readiness presented in the main manuscript (Figure 1). It is offered to support readers in tracing the relationship between the qualitative findings and the integrated interpretive account.

| **Theme** | **Key Empirical Insight** | **Mechanism Identified** | **Conceptual Contribution to Model** |
| --- | --- | --- | --- |
| **Theme 1: Discharge as Anticipatory Nursing Work** | Nurses described discharge preparation as beginning at admission and sustained through ongoing assessment, coordination and problem-solving embedded in everyday nursing practice. Structural tools (checklists, booklets) scaffolded this work in Australia; iterative interprofessional engagement characterised it in the UK. | Continuous clinical surveillance, prospective planning and anticipatory coordination of practical, functional and social arrangements across the full admission trajectory. | Establishes discharge readiness as a temporal process produced across admission, not confirmed at its end. Nurses' anticipatory orientation constitutes the foundational layer of discharge labour. |
| **Theme 2: Discharge as Collective Accomplishment** | Discharge readiness emerged through ongoing interaction between nurses, doctors, pharmacists and allied health professionals, each contributing discipline-specific expertise. Nurses anchored these negotiations through continuous patient contact; pharmacists (AUS) and allied health (UK) shaped feasibility and functional readiness. | Distributed clinical expertise and interprofessional negotiation, with nursing coordination connecting and initiating contributions across disciplines. Professional hierarchy mediated whose voice carried weight. | Demonstrates discharge readiness as co-constructed through negotiation, not determined by any single professional group. Nurses' coordinative role is analytically central, even where formal authority over the discharge decision rests elsewhere. |
| **Theme 3: Discharge Documentation as a Site of Tension** | Clinicians described documentation as essential to continuity of care but also burdensome and poorly aligned with the | Administrative requirements structuring how clinicians allocated time, what discharge work was made | Reveals documentation as an externally imposed organisational mechanism that determines what |
|  | practical communication needs of patients and receiving providers. Documentation competed with time for patient education and relational discharge work. | visible, and what constituted institutional completion of discharge. Documentation functioned as an organisational mechanism shaping clinical priorities. | discharge work counts, what remains invisible, and how the tension between administrative and clinical logics is resolved in everyday practice. |
| **Theme 4: Discharge Within System and Organisational Pressures** | Discharge timing was shaped by bed pressure, staffing levels, weekend service gaps, community capacity and geographic access constraints. Australian clinicians navigated pharmacy and transport access at the system periphery; UK clinicians described throughput pressure and community capacity failures at the system centre. | Institutional priorities and structural conditions interacting with clinical readiness to shape when discharge was feasible. The rate-limiting step on discharge was frequently beyond CCU control. | Positions discharge readiness as fundamentally contingent, not a property of the patient that clinicians confirm, but a relational achievement produced through alignment between patient state and surrounding system capacity. Anticipatory work is necessary but not sufficient without system conditions that make discharge achievable. |

### Integrative Interpretation

The four themes demonstrate that discharge readiness in coronary care units emerges through the interaction of clinical, relational and organisational processes rather than through a single clinical decision. Anticipatory nursing work establishes the foundational layer of discharge labour, with nurses continuously monitoring patient progress and maintaining a longitudinal view of readiness across the admission. Interprofessional negotiation determines whether patients are considered sufficiently stable, supported and practically resourced for discharge, with nursing input anchoring discussions that draw on distributed disciplinary expertise. Documentation practices structure how discharge information is communicated and what counts institutionally as completed discharge work, while broader organisational pressures shape the timing and feasibility of discharge decisions independently of clinical readiness.

Within the conceptual model proposed in the main manuscript, discharge readiness can be understood as the emergent outcome of three interacting domains:

Clinical coordination work undertaken by nurses and allied health professionals across the admission

Interprofessional negotiation of readiness between clinical disciplines, anchored but not owned by nursing

Organisational and system-level conditions that shape the timing and outer limits of what anticipatory work can achieve Together these mechanisms explain how discharge decisions are produced within complex healthcare environments, extending existing conceptualisations of transitional care by highlighting the distributed, system-embedded and often-invisible nature of discharge readiness.

### Supplementary File 4

Anticipatory Discharge Work in Coronary Care: A Practice-Informed Clinician Checklist

#### About this checklist

This checklist is offered as an illustrative translation of the qualitative findings from this study into a structured practice tool. Items are organised around the four empirical themes generated through analysis of clinician accounts in coronary care units (CCUs) in Australia and the United Kingdom: anticipatory nursing work; interprofessional negotiation; documentation as organisational mechanism; and system and organisational conditions. The checklist is intended as a flexible, practice-informed prompt to support anticipatory and interprofessional discharge work across the admission. It is not a validated intervention, a prescriptive protocol, or a standalone instrument for measuring discharge readiness. Items reflect the patterns identified in the qualitative findings, supplemented where appropriate by established cardiac discharge guidance and recognised communication and coordination practices. As the checklist is derived from qualitative findings rather than evaluated empirically, further research is required to test its feasibility, acceptability and impact in different clinical contexts.

### How to use this checklist

The checklist is organised into four sections, one per empirical theme. Each section opens with a short framing note that links the items to what participants described in the analysis. The items are not intended to be completed sequentially or in full for every patient; they are prompts to support attention to the dimensions of discharge work the findings identified, adaptable to local service pathways and resource conditions. Local services may wish to integrate selected items into existing discharge documentation, multidisciplinary review processes, or audit tools.

### Alignment with existing guidance

Where relevant, the checklist is consistent with principles reflected in established cardiac discharge guidance and policy frameworks, including NICE guidance on transitions of care between inpatient and community settings (NICE, 2015), the Australian Commission on Safety and Quality in Health Care National Safety and Quality Health Service Standards (ACSQHC, 2017, 2020), and the WHO Framework on Integrated People-Centred Health Services (WHO, 2016). It is intended to complement rather than replace local discharge protocols, cardiac care pathways or organisational policy.

### Reference

Rashidi A, Dunham M, Kinley E, Bueser T, Glass C, Jones I, Makokha M, Whitehead L, Newson L. Anticipatory nursing work and the co-construction of discharge readiness in coronary care units: A cross-national qualitative study. Supplementary File Discharge Checklist [Journal name, year, to be completed at publication].

### IP

This checklist was conceptualised and developed by Professor Newson & Dr Kinley at Liverpool John Moores University, as part of the research reported in this manuscript. It should be cited using the associated manuscript/preprint. Copyright and reuse are governed by the licence applied to the preprint and any applicable institutional rights.

### Clinician Practice Checklist

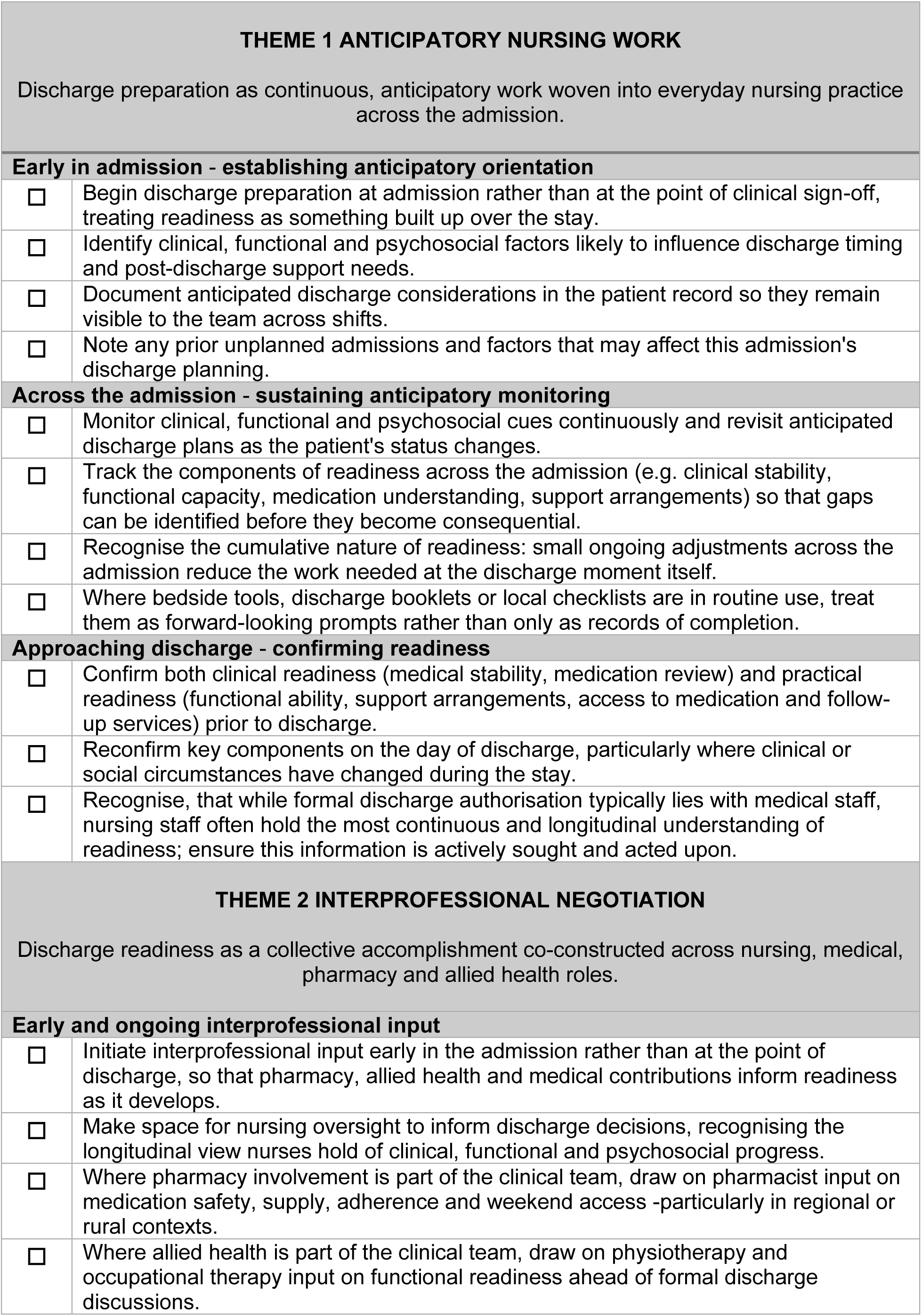
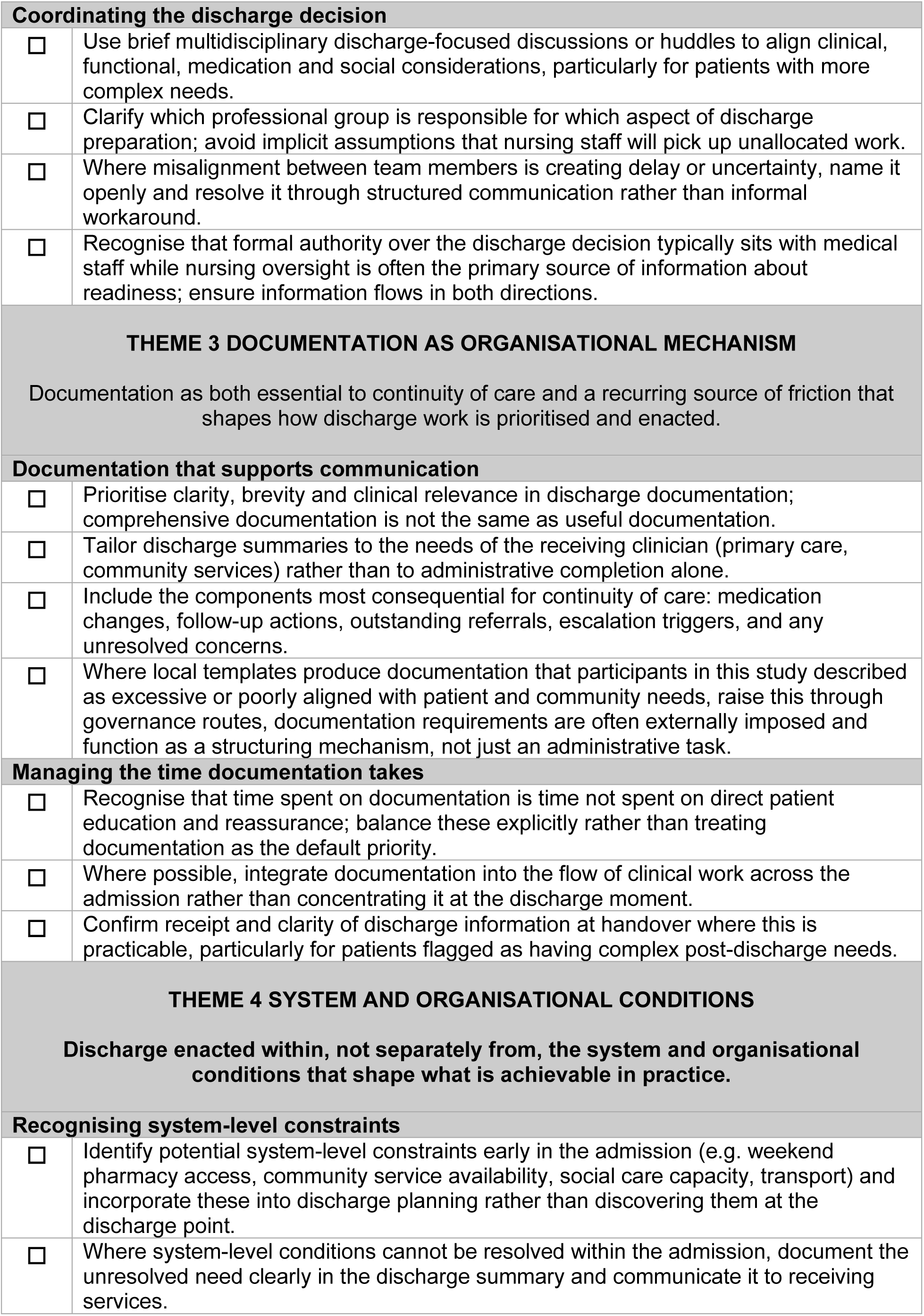
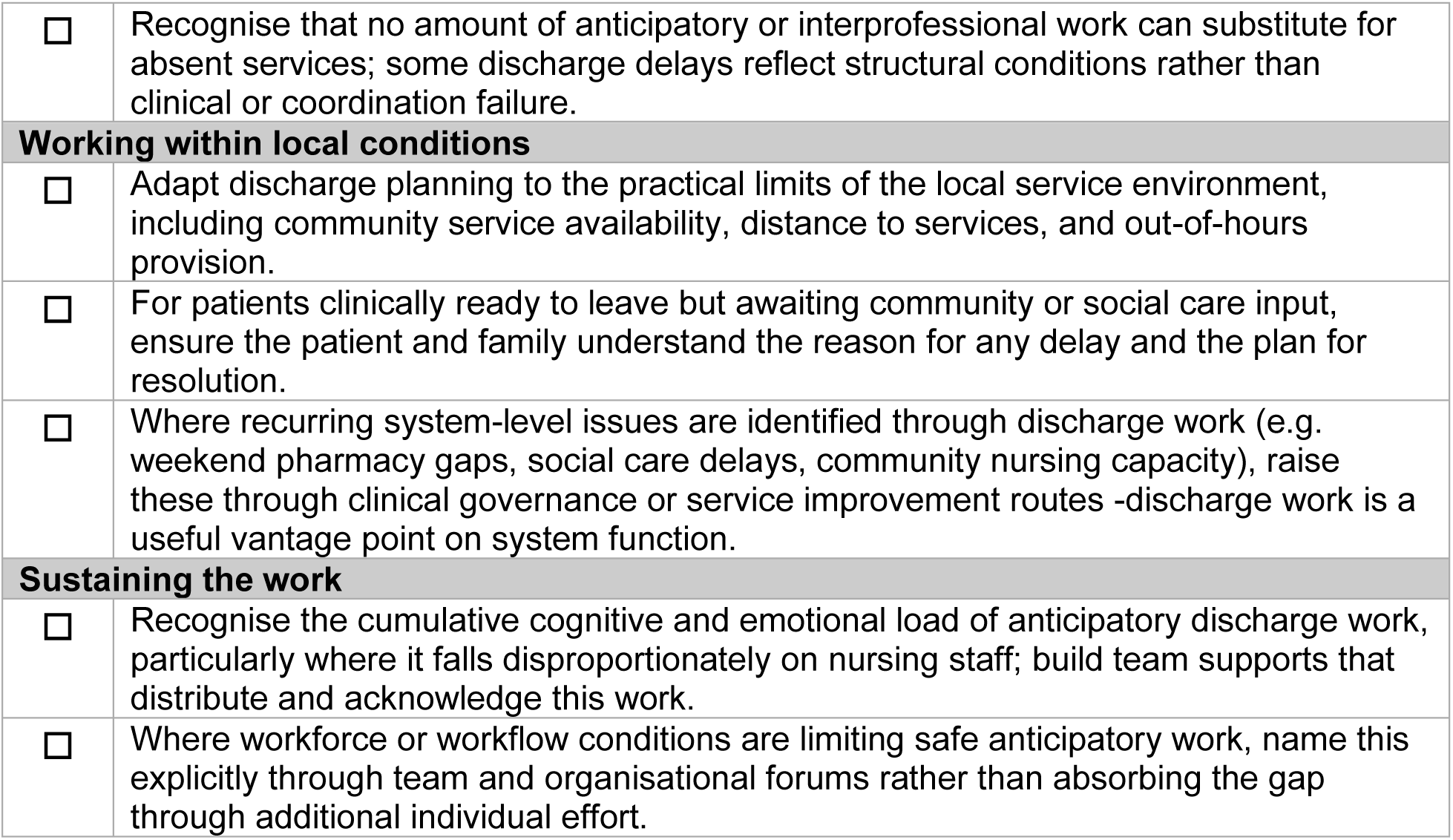

### Implementation notes

These notes outline optional approaches to support implementation of the checklist in routine practice. They are illustrative rather than prescriptive and are intended to support local adaptation rather than to specify particular tools, products or pathways. Local services should integrate the checklist with existing cardiac discharge protocols, organisational policy and local service pathways.

### Anticipatory nursing work

- Local discharge documentation can be reviewed for whether it functions as a forward-looking tool (prompting attention to upcoming readiness needs) or only as a backward-looking record (capturing what has been completed). Both are useful; the anticipatory function is the one most directly supported by the findings.
- Routine team discussion of anticipated discharge needs at admission and across the stay, even brief, can help make the cumulative nature of readiness visible.
- Recognising and naming nursing coordinative work explicitly in clinical handover and documentation supports its visibility to the wider team.

### Interprofessional negotiation

- Brief multidisciplinary discharge-focused discussions (sometimes used as huddles) typically take five to ten minutes and can clarify roles, medication considerations, follow-up actions and risks. Local services may integrate these into existing ward rounds or board rounds.
- Standardised communication formats (e.g. SBAR or equivalent) can support consistent information transfer across shifts and between disciplines, particularly where team composition varies.
- Where pharmacy or allied health involvement is variable across shifts or weekends, discharge planning may need to anticipate these gaps rather than assume continuous availability.

### Documentation as organisational mechanism

- Local services may wish to review discharge summary templates for clarity, length and alignment with the needs of receiving primary care and community providers.
- Where electronic health record systems generate lengthy or template-driven summaries, a brief targeted summary highlighting the most consequential information for continuity of care may complement rather than replace the formal record.
- Confirming receipt of discharge information by primary care or community services, where practicable, can support continuity for patients with complex post-discharge needs.

### System and organisational conditions

- Routine recording of system-level reasons for discharge delay (e.g. medication supply, community service capacity, social care availability) supports clinical governance visibility of structural conditions affecting discharge.
- Where particular system-level constraints recur (weekend pharmacy access in regional contexts; community nursing or social care capacity; transport limitations), these may benefit from organisational or system-level attention rather than case-by-case workaround.
- Workforce planning that explicitly recognises and resources the coordinative dimensions of nursing discharge work support the conditions through which anticipatory work is produced.

### Limitations of this checklist

This checklist is derived from qualitative findings in coronary care units in two healthcare systems. It has not been empirically evaluated, validated as an instrument, or tested against patient-level outcomes. It does not replace clinical judgement, local protocols, or formal discharge processes. Items reflect what clinicians described as shaping discharge work rather than the experiences of patients or families, who were not part of this study. The checklist should be adapted to local service conditions and used in conjunction with established cardiac discharge guidance.

